# A large-scale genome-wide association meta-analysis for nevus count provides direct insights into the genetics of melanoma

**DOI:** 10.1101/2025.02.25.25322831

**Authors:** G J M S R Jayasinghe, Gu Zhu, Nirmala Pandeya, Catherine M Olsen, Nicholas G Martin, Penelope A Lind, Sarah E Medland, Scott D Gordon, Santiago D Torres, Gareth Lingham, Samantha S Y Lee, Tamar Nijsten, Manfred Kayser, Luba M. Pardo, Grant W Montgomery, Nicholas K Hayward, Jane M Palmer, David J Hunter, Jiali Han, Alex W Hewitt, Mario Falchi, D Timothy Bishop, Kevin M Brown, Veronique Bataille, David A Mackey, Mark M Iles, David C Whiteman, David L Duffy, Stuart MacGregor, Matthew H Law

**Author notes:** **Correspondence:** Matthew H Law, G J M S R Jayasinghe.

## Abstract

A greater understanding of the biology of nevi will provide insights into the etiology of melanoma. Our large-scale meta-analysis of 14 nevus genome-wide association study (GWAS) included 85,967 individuals of European ancestry. We identified 29 nevus-associated loci (p < 5×10^−8^), of which 24 were not previously reported in a GWAS conducted for nevus count alone. We further identified 255 candidate genes for nevus loci, including *SIKE1* which is involved in immune response regulation. This is of interest because immune response regulation influences the formation of nevi and melanoma susceptibility. Gene-set enrichment analyses prioritised immune response-related pathways and cancers that do not have a pigmentation component (e.g. breast, prostate, and glioma). This suggests that the biology underlying nevus count captures risk pathways beyond pigmentation that are relevant to melanoma. A nevus polygenic risk score explains 5% of the variance in nevus count, indicating its potential to enhance melanoma risk prediction.

## Introduction

Melanocytic nevi are small, pigmented benign skin tumours composed of melanocytes. Melanoma is the deadliest form of skin cancer and arises from the malignant transformation of melanocytes. Melanoma can develop from a pre-existing nevus or appear as a new lesion on the skin^1,2^. A high melanocytic nevus count is the strongest single risk factor for melanoma^3^. Therefore, understanding the biology of nevi may also provide insights into the etiology of melanoma.

Nevus count is a highly heritable trait with heritability estimates from twins ranging from 60% to 70% (Lee et al., 2016). Previous genome-wide association studies (GWAS) for nevus count (N > 52,000) have identified five significant loci^4^. These loci were also associated with melanoma risk^4^, implying both traits have a shared genetic risk component. Although combined analyses of both melanoma and nevus count have identified additional loci that may be pleiotropically associated with both traits^4,5^, given the high heritability of nevus count, many causal genetic variants remain to be discovered.

In addition to providing insights into cutaneous melanoma, nevus count genetics can improve our understanding of rarer forms of melanoma. Uveal melanoma is a rare but deadly form of melanoma originating from ocular melanocytes. Atypical nevi and iris nevi are risk factors for uveal melanoma^6–8^, suggesting that the findings from a cutaneous nevus GWAS can be applied to risk factors for uveal melanoma.

To identify further genetic variants associated with nevus count, we meta-analysed 14 GWASs of nevus count, comprising over 85,000 individuals, including two previously unpublished datasets. We also report on the application of post-GWAS functional analyses to identify those genes most strongly involved in nevus biology and to identify the pathways underlying these genes. Finally, we generated polygenic scores for nevus count and tested their ability to predict nevus count in adults and to predict iris nevus count.

## Results

### Nevus GWAS meta-analysis

We initially estimated the degree to which genotyped SNPs could capture variation in nevus count within the QSkin I cohort (*n* = 15,346). The SNP-based heritability of nevus count estimated using GCTA-GREML was 0.22 (SE = 0.03).

We then performed a large-scale GWAS meta-analysis of nevus count with a total sample size of 85,965 participants. The genomic inflation factor λ and LDSC intercept for the GWAS meta-analysis suggested polygenic inheritance (λ = 1.068, intercept = 1.014, ratio = 0.119). In the nevus meta-analysis, 41 independent genome-wide significant SNPs were assigned to 29 genomic loci (Figure 1). Of these 29 loci, 24 have not previously been reported in a GWAS conducted solely for nevus count (Table 1). Nineteen of these 24 loci have been previously identified as associated with both nevus count and melanoma in joint analyses^4,5^. Among the remaining five loci after 19/24, the regions near *MAP3K1* on chromosome 5 and around *TPCN2* on chromosome 11 have been also associated with melanoma and hair colour^5^. LDSC regression analysis estimated the proportion of the variance in nevus count captured by the GWAS SNPs in the current GWAS meta-analysis at 0.06 (SE = 0.01); while this is expected to be an underestimate relative to GCTA-GREML as it uses fewer SNPs and summary data, this estimate of variance is two-times higher than that of the previous nevus GWAS meta-analysis^4^

**Figure 1.**
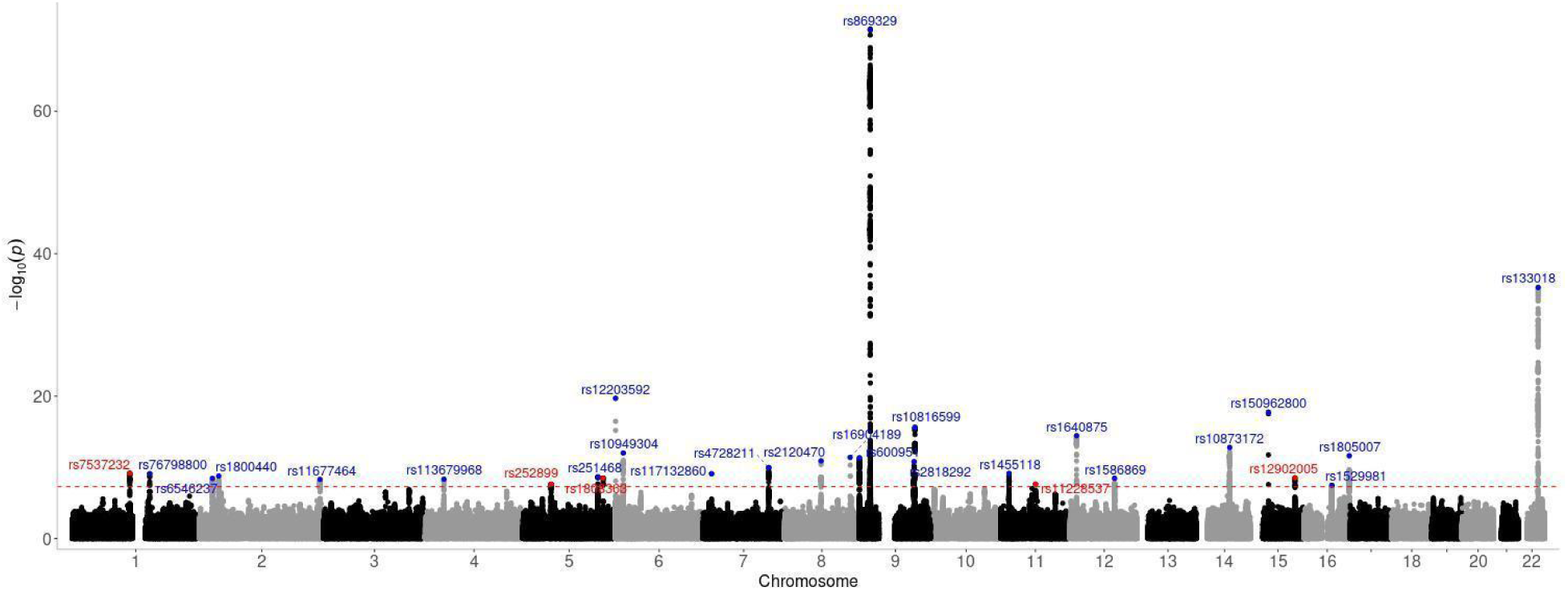
Manhattan plot of the GWAS of nevus count. Each dot represents a variant, the x-axis shows the chromosome location of each SNP and the y-axis shows its association with the nevus count. The red dashed line shows the genome-wide significance threshold (P < 5×10^−8^; −log_10_P < 7.3). Twenty-nine genomic risk loci reached the genome-wide threshold. Novel loci are represented in the plot as red dots, whereas loci reported in previous nevus GWASs, nevus/melanoma or nevus/pigmentation combined GWASs are presented in blue.

**Table 1:** Top significant SNPs at each locus identified by the GWAS meta-analysis for nevus count.

| rsID | chr | pos | EA/NEA | Freq | P | Zscore | Genes |
| --- | --- | --- | --- | --- | --- | --- | --- |
| rs7537232* | 1 | 115284056 | T/G | 0.26 | 6.80x10<sup>-10</sup> | 6.17 | *BCAS2, SIKE1* |
| rs76798800 | 1 | 154994978 | T/G | 0.247 | 7.81x10<sup>-10</sup> | 6.15 | *PBXIP1, ADAM15* |
| rs6546237 | 2 | 25749376 | T/C | 0.782 | 3.67x10<sup>-9</sup> | -5.90 | *DTNB* |
| rs1800440 | 2 | 38298139 | T/C | 0.824 | 1.72x10<sup>-9</sup> | 6.02 | *CYP1B1* |
| rs11677464 | 2 | 240065356 | A/G | 0.904 | 4.94x10<sup>-9</sup> | -5.85 | *HDAC4* |
| rs113679968 | 4 | 37439840 | A/G | 0.147 | 4.68x10<sup>-9</sup> | -5.86 | *C4orf19* |
| rs252899* | 5 | 56188569 | C/G | 0.81 | 2.37x10<sup>-8</sup> | -5.58 | *MAP3K1* |
| rs251468 | 5 | 149194485 | T/C | 0.28 | 2.17x10<sup>-9</sup> | -5.99 | *PPARGC1B* |
| rs1808363* | 5 | 159888522 | A/G | 0.525 | 3.22x10<sup>-9</sup> | -5.92 | *PTTG1* |
| *rs12203592* | 6 | *396321* | *T/C* | *0.131* | *2.02x10<sup>-20</sup>* | *-9.26* | *IRF4* |
| rs10949304 | 6 | 15503696 | C/G | 0.451 | 9.70x10<sup>-13</sup> | 7.14 | *JARID2, DTNBP1* |
| rs117132860 | 7 | 17134708 | A/G | 0.019 | 7.88x10<sup>-10</sup> | 6.15 | *AHR* |
| rs4728211 | 7 | 130876086 | T/C | 0.13 | 1.06x10<sup>-10</sup> | 6.46 | *MKLN1* |
| rs2120470 | 8 | 72863438 | T/G | 0.366 | 1.26x10<sup>-11</sup> | -6.77 | *MSC* |
| rs16904189 | 8 | 131025191 | T/C | 0.959 | 4.01x10<sup>-12</sup> | 6.94 | *GSDMC* |
| *rs600951* | 9 | *224742* | *A/G* | *0.463* | *4.74x10<sup>-12</sup>* | *6.91* | *DOCK8* |
| *rs869329* | 9 | *21804693* | *A/G* | *0.477* | *3.2x10<sup>-72</sup>* | *17.97* | *MTAP, CDKN2A* |
| rs2818292 | 9 | 109011615 | A/C | 0.738 | 1.63x10<sup>-11</sup> | -6.74 | *TMEM38B* |
| rs10816599 | 9 | 110716721 | C/G | 0.332 | 2.44x10<sup>-16</sup> | -8.20 | *KLF4* |
| rs1455118 | 11 | 16221683 | T/C | 0.516 | 7.40x10<sup>-10</sup> | 6.16 | *SOX6* |
| rs11228537* | 11 | 68927358 | A/G | 0.886 | 2.31x10<sup>-8</sup> | 5.59 | *TPCN2* |
| rs1640875 | 12 | 13069524 | A/T | 0.563 | 3.62x10<sup>-15</sup> | -7.87 | *GPRC5D* |
| *rs1586869* | 12 | *88980649* | *A/G* | *0.827* | *3.5x10<sup>-9</sup>* | *5.91* | *KITLG* |
| rs10873172 | 14 | 64390030 | C/G | 0.71 | 1.63x10<sup>-13</sup> | -7.38 | *SYNE2* |
| rs150962800 | 15 | 33260973 | T/C | 0.017 | 1.85x10<sup>-18</sup> | 8.77 | *FMN1* |
| rs12902005* | 15 | 86091174 | A/G | 0.137 | 2.90x10<sup>-9</sup> | 5.94 | *AKAP13, KLHL25* |
| rs1529981 | 16 | 55323241 | T/G | 0.237 | 3.4x10<sup>-8</sup> | 5.52 | *MMP2* |
| rs1805007 | 16 | 89986117 | T/C | 0.063 | 2.40x10<sup>-12</sup> | -7.01 | *MC1R* |
| *rs133018* | 22 | *38572761* | *A/G* | *0.481* | *5.36x10<sup>-36</sup>* | *-12.53* | *PLA2G6* |

Abbreviations: chr - chromosome, pos - position (hg37), EA - effect allele, NEA - non-effect allele, Freq - Effect allele frequency. Loci reported in a previous GWAS conducted solely for nevus count are italicised. Gene prioritises the functional target if known previously followed by based on the genes prioritised by all three mapping approaches or the nearest protein-coding gene. \* Novel loci not reported in previous nevus GWASs, nevus/melanoma or nevus/pigmentation combined GWASs

### Pleiotropy of nevus loci

To determine whether the significant nevus loci were pleiotropic with melanoma, a pairwise GWAS was conducted by integrating our results with a previously published melanoma GWAS ^5^. This approach assigns a posterior probability of association (PPA), which sum to 1, to four models; models 1 and 2 suggest the gene region is only associated with trait 1 or 2 respectively; model 3 suggests pleiotropy and model 4 tests an independent association^9^ (see Methods). Twenty-eight of 29 nevus loci exhibited strong evidence of pleiotropy regions for both melanoma and nevus count (model 3 PPA > 0.95) (Supplementary Table 7). The 29th locus, near TPCN*2* on chromosome 11 has the highest PPA for model 4 (independence; PPA 0.76) but as no model reached the threshold (> 0.95) we are unable to reach strong conclusions about the role of this locus.

#### Functional mapping

We subsequently used FUMA to identify potential functional links between nevus-associated variants and candidate genes at genome-wide significant loci (Methods). Using three mapping approaches (positional, eQTL and chromatin interaction mapping) 248 genes were prioritised across the nevus count loci (Supplementary Table 2). These prioritised genes included previously reported nevus (e.g. *KITLG, MTAP/CDKN2A* and *IRF4*) or melanoma-related genes (e.g. *MC1R*, *TMEM38B* and *DCST2*)^4,5,10^. Of 248 genes, 26 mapped on 14 of 29 nevus-associated genomic loci were identified by positional, eQTL and chromatin interaction mapping. Interestingly, of these 26 genes, we identified candidate genes for 2/3 novel genomic regions that have not previously been reported in a nevus GWAS, nevus/melanoma combined analysis or a nevus/pigmentation combined analysis, marked by rs7537232 (*BCAS2* and *SIKE1)* and rs12902005 (*AKAP13* and *KLHL25*) as indicated in Table 1. In addition, *PTTG1* was identified as the candidate gene for the remaining nevus-associated novel locus on chromosome 5 through the positional and eQTL mapping.

#### Gene-based analyses

MAGMA gene-based analysis evaluates candidate genes for association with a trait by combining the local association test statistics for SNPs within a gene region. This approach can identify additional significant genetic loci that may not reach genome-wide significance in single SNP tests. MAGMA identified 33 genome-wide significant genes (P < 2.64×10^−6^) at 18 genomic loci; three were in addition to those found in our GWAS meta-analysis (Figure 2 and Supplementary Table 3). Of these 33 genes, 28 overlapped with those prioritised by functional mapping approaches and included well-known nevus-, melanoma-, or pigmentation-associated genes providing further evidence of their shared genetics. The three additional loci identified by MAGMA included a novel locus not previously identified for nevus count or melanoma on chromosome 22 (*C220rf26*), a locus previously found in nevus/melanoma combined analysis on chromosome 10 (*FAM208B*)^4^, and a locus previously reported for melanoma on chromosome 3 (*MYNN*)^10^. Interestingly, through MAGMA gene analysis we found *NRAS* as a significant candidate gene for nevi at the nevus risk locus marked by rs7537232.

**Figure 2.**
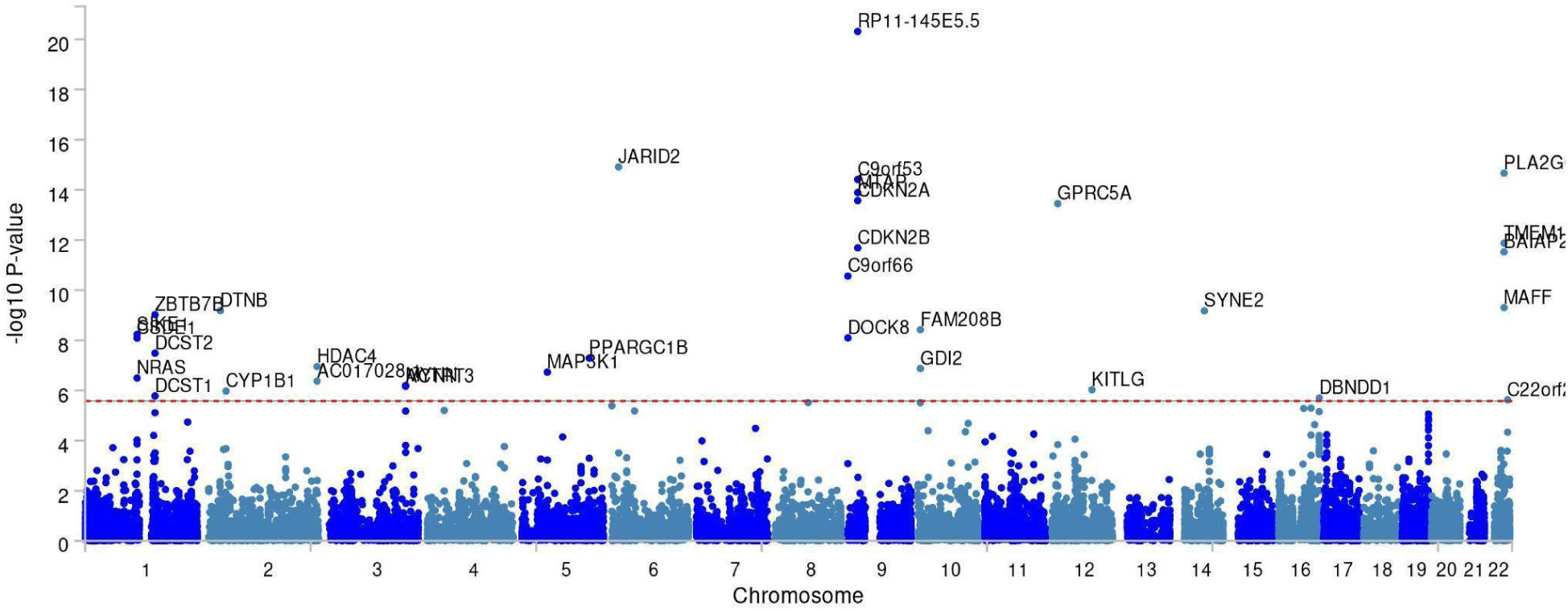
MAGMA gene-based analysis identified 33 genes associated with nevus count. Each dot represents a gene, the x-axis shows the chromosome location of each gene and the y-axis shows its association with nevus count. The red dashed line shows the genome-wide significance threshold (P < (0.05/19,055) 2.6×10^−6^; −log10P > 5.58).

### Transcriptome-wide association study (TWAS)

To expand on our gene-mapping work, we carried out TWAS to nominate plausible functional genes linked to nevus count by testing for significant correlation with gene expression. We analysed gene expression data from three skin-related tissue-based cis-eQTL datasets (sun-exposed, not-sun-exposed, and fibroblasts) available in GTEx (see Methods), melanocyte-specific eQTL data^5^ and an additional well-powered but potentially less tissue-relevant whole blood dataset from the eQTLGen consortium. After applying multiple-testing corrections the five TWAS analyses identified a range of promising candidate genes. Across all TWAS analyses, candidate genes were identified at 12 genome-wide significant nevus loci and two additional loci on chromosome 10 (*LRMDA* and *GDI2*) (Supplementary Table 6.1 and 6.2). The region near *GDI2* (Supplementary Table 6.1) overlaps with the locus; *FAM208B* found through MAGMA gene-based analysis (Supplementary Table 3).

### Pathway analyses

In the MAGMA gene-set analysis, three gene sets involved in the regulation of the microtubule cytoskeleton during radial glia-guided migration in the cerebral cortex, development of astrocytoma in patients with Neurofibromatosis Type 1 (NF1) and regulation of the catalytic activity (i.e. any process that modulates the activity of enzyme), were found to be significant after Bonferroni correction (Supplementary Table 4).

In addition to using the MAGMA data, we conducted gene-set enrichment tests using the genes prioritised by any of the three gene mapping approaches in FUMA. Notably, prioritised genes were significantly enriched in gene sets related to cancers (e.g. breast, prostate and glioma cancer) that do not have a pigmentation component. As expected, gene-sets previously reported for nevus, melanoma and pigmentation were also enriched (Supplementary Table 5). Fifteen genes (including *NRAS, THBS3, IFNA16, CCND1* and *KITLG*) were enriched in the critical cell survival, growth and apoptosis P13K/AKT signaling pathway (adjusted P = 1.68e-5). In addition, we found significant enrichments in several immune response-related pathways. The top two pathways were the type 1 interferon receptor binding pathway and RIG-I-like (Retinoic acid-inducible gene I) receptor signaling pathway (Supplementary Table 5). Further, 28 prioritised genes were significantly overrepresented in the gene set involved in response to the organic cyclic compounds. Of transcriptional factor target gene sets, four gene sets showed significant enrichment signals, of which the most significant was the target gene set regulated by transcription factor PRDM4 (PR domain zinc finger protein 4), which is involved in cell proliferation and tumourigenesis^11,12^.

### Polygenic risk score analysis for nevi

We built a nevus PRS using SBayesRC applied to a meta-analysis of 13 out of the 14 nevus GWASs (excluding QSkin II) (see Methods). We tested the nevus PRS on the left-out QSkin II data and the model including nevus PRS explained 5% (R^2^ = 0.05) of the variation and a beta coefficient per (standard deviation) SD of the nevus PRS was 0.19 (SE = 0.009, P < 4.94e^−324^, *n* = 6,608). Further, we applied the same nevus PRS to the Kidskin Young Adult Myopia Study (KYAMS) cohort and the nevus PRS model performance was closely similar to that in QSKIN II where R^2^ was 4% and beta was 0.20 (SE = 0.060, P = 0.0009, *n* = 265). Finally, ocular nevi are a risk factor for the rare and deadly uveal melanoma. We tested whether our PRS for cutaneous nevi was associated with iris nevi scored in the QIMR Brisbane Twin Nevus Study (BTNS) cohort. After generating a new PRS without the BTNS cohort, this PRS was weak but significantly associated with iris nevus count (R^2^ = 0.0018, beta per SD of the PRS = 0.03, SE = 0.016, P = 0.048, *n* = 2,607)

## Discussion

In this study, we performed the largest nevus GWAS meta-analysis to date identifying 29 genome-wide significant genomic loci. Of these, 24 had not previously been reported in a prior GWAS conducted solely for nevus count^4^. Prior analyses have leveraged the strong genetic overlap between melanoma and nevus count to test for regions of the genome pleiotropically associated with both traits. Promisingly most of the loci newly identified for nevus count alone (19/24) were significant in these combined analyses^4,5^. Of the remaining five loci, two (*TPCN2* and *MAP3K1*) had been identified in a joint analysis of melanoma and hair colour^5^ while the other three loci (*BCAS2*/*SIKE1*, *AKAP13*/*KLHL25* and *PTTG1*) are completely novel and had not been identified in previous GWASs conducted for nevus count, melanoma or pigmentation^5^. *BCAS2,* known to be amplified and overexpressed in breast cancer cell lines^13,14^ may point to a potential role in cell proliferation that may contribute to nevus development. *SIKE1* is involved in regulating immune responses particularly those triggered by viruses and TLR3 (Toll-like receptor 3)^15^. *SIKE1* may influence nevi growth by potentially affecting the immune-mediated control of melanocyte proliferation. *AKAP13*, identified as a tumour suppressor cooperating with *PTEN* in prostate cancer^16^ may similarly influence nevus formation and melanoma risk. Finally, silencing of *PTTG1* in melanoma cell lines impaired cell proliferation and invasiveness^17,18^.

In addition, MAGMA gene-based analysis and TWAS identified four additional loci that were not genome-wide significant in the meta-analysis, including novel loci on chromosome 10 (*LRMDA,* also known as *C10orf11*) and chromosome 22 (*C220rf26*) for which function is unclear. A melanocyte-differentiation gene, *LRMDA* when mutated, causes autosomal recessive albinism^19,20^. *LRMDA* is involved in melanocyte differentiation and melanosome maturation^20^, which could provide insights into its genetic influence on nevus formation and possibly melanoma risk. MAGMA gene-based analysis identified *NRAS* as a significant candidate gene for the novel locus marked by rs7537232. Of note, *NRAS* codon 12/13 or 61 mutation is found in all (∼15%) *BRAF*-mutant-negative acquired nevi^21^. *NRAS* is the second most common driver mutation in melanoma and is frequent in congenital melanocytic nevi^22^.

As expected, when we performed our joint analyses of nevus count and melanoma, we found that the nevus-associated loci were pleiotropically associated with both traits; only one locus (*TPCN2*) did not strongly support any of the models. Based on the nevus count vs melanoma effect size plot for the identified 29 significant nevus loci (Supplementary Figure 19), the *MC1R* variant (rs1805007) shows an inverse relationship; the allele that reduces nevus count increases the risk of melanoma. This may represent a true antagonistic biological effect on the two traits or may arise from the difficulty in identifying and counting nevi in individuals with red hair and very low skin pigmentation. Further studies using objective nevus detection methods (e.g. imaging technologies) and exploring the effect of *MC1R* variants on nevus count are needed to clarify this potentially inverse relationship.

We also performed *in-silico* assays to determine likely candidate genes at identified loci. According to gene-set analysis in FUMA, prioritised genes (e.g. *PLA2G6, CDKN2A, CCND1, GPRC5A* and *SYNE2*) from three gene mapping approaches (positional, eQTL and chromatin interaction mapping) showed enrichment in known pigmentation-related traits such as hair colour and tanning response (e.g. *MAP3K1, PLK2, IRF4, EXOC2, DOCK8, TMEM38B* and*, TPCN2*), reinforcing the shared genetics of nevus development and melanoma^4,5,23,24^. As well as known pathways, prioritised genes showed enrichments in several immune response-related pathways (e.g. the biological process involved in regulating peptidyl-serine phosphorylation of STAT protein, Supplementary Table 5) that may be critical in nevus formation and progression to melanoma, as immune dysregulation can drive tumour development through altered cell growth and inflammation.

Further, we identified gene set enrichments in cancers that do not have a pigmentation component. This finding underscores the significance of analysing nevus count, as it can reveal biological pathways relevant to melanoma that fall outside the known risk pathways (i.e. skin colour). In addition, MAGMA gene-set analysis, as well as FUMA gene-set analysis, revealed that candidate genes are significantly overrepresented especially in brain tumours, particularly glioblastomas and astrocytomas with neurofibromatosis type 1 (Supplementary Tables 4 and 5). A connection between melanoma and these brain tumours has been observed in families with multiple melanomas and atypical mole syndrome ^25^. Since melanocytes originate from the neural crest^26^, they share a developmental lineage with certain brain cells, which may explain this association. In addition, there is an association between nevi and neurofibromatosis type 1^27^. These facts show that studying nevi can reveal biological pathways intersecting with brain function and neurological diseases. This shared origin positions nevi as a valuable focus for exploring connections between melanoma, brain biology, and related disorders.

In addition, the biological pathway associated with responding to organic compounds suggests that the genes enriched in this set likely play a significant role in responding to DNA damage, particularly from ultraviolet (UV) exposure. For instance, *AHR* and *CYP1B1* were included in this gene set. *AHR* plays a role in melanoma risk through its interaction with UV exposure,^28^. *CYP1B1*, an *AHR* target, has a risk variant that impacts melanoma susceptibility^28^ and has been found to interact with environmental exposures (such as cook stove smoke) linked to increased cancer risk^29^. Accordingly, the candidate genes that are enriched in this pathway may offer potential target therapeutics that enhance DNA repair or mitigate UV-induced damage.

We constructed a PRS for nevi that showed promising predictive power in two independent adulthood cohorts: the older adulthood cohort QSkin II (R^2^ = 0.05) and the young adulthood cohort KYAMS (R^2^ = 0.04), indicating its effectiveness in capturing genetic variance associated with nevus development.

Iris nevi and atypical cutaneous nevi are risk factors for uveal melanoma, a rare but deadly form of melanoma in the eye. We thus explored if a genetic predisposition to cutaneous nevi formation also contributes to the development of iris nevi. The cutaneous nevus PRS yielded a lower (but statistically significant) predictive power (R^2^ = 0.0018) for iris nevi compared to what we observed for cutaneous nevi. This may be because the overlap in genetic factors influencing iris nevi is incomplete. Future research should explore these differences further and investigate whether modifications to the nevus PRS can enhance its predictive ability for iris nevi and eventually uveal melanoma.

The strengths and limitations of our study should be considered in interpreting the findings. Our study was limited to individuals of European ancestry; hence it may not be readily extendable to other populations. Our analysis was limited to individuals who were melanoma-free at the time of the last follow-up. While we cannot rule out that some may have subsequently been diagnosed with melanoma, numbers would be low and unlikely to bias our results. Our study presents notable strengths. First, the large sample size compared to the previous nevus GWASs provided robust statistical power to identify several novel loci associated with nevus count. Second, through applying several post-GWAS analyses, we identified candidate genes in tissues; melanocytes and skin, that are relevant to the nevus and melanoma biology. Third, owing to the improvement in statistical power, the out-of-sample prediction ability of our nevus PRS is promising. This demonstrates the potential utility of this nevus PRS in identifying genetic predispositions that span both cutaneous and eye nevi which may enhance the risk assessment of deadly cutaneous melanoma and uveal melanoma.

In summary, our GWAS, the largest to date, identified novel loci solely associated with nevus count as well as biological pathways unrelated to pigmentation, which is promising for future research towards therapeutic targets for melanoma. Through our study, we constructed a nevus PRS with promising predictive ability and in future, by integrating the nevus PRS into melanoma risk prediction models, we anticipate improved accuracy in melanoma risk assessment, leveraging shared genetic factors between nevus count and melanoma susceptibility for more precise risk stratification.

## Online Methods

### GWAS cohorts

We used GWAS summary statistics for nevus count from 11 studies that were reported in a previously published nevus GWAS meta-analysis of 52,236 individuals of European ancestry^4^. These studies were combined with new data from the QSkin Sun and Health Study cohort^30^ Phase I (QSkin I, *n* = 15,346) and Phase II (QSkin II, *n* = 6,608), and the Australian Genetics of Depression Study (AGDS, *n* = 11,775) ^31^. A subset of QSkin I (*n* = 12,930) with nevus count that was non-overlapping and unrelated (identical-by-descent score < 0.15) to the QSkin melanoma case-control set was included in a previous pleiotropic GWAS analysis with melanoma^5^. However, in the current GWAS of QSkin I, we only excluded QSkin melanoma cases, increasing the number of available samples for this analysis of nevus loci (*n* = 15,346). Details of each cohort including genotyping, quality control, imputation, and phenotypic descriptions, are provided in Supplementary Notes S1.1 and S1.2. All study participants over 18 years provided written informed consent; for those under 18 years consent was obtained from parents and full details of ethics approvals (QSkin I: P1309 and P2034, QSkin II: P3434, AGDS: P2118) have been reported elsewhere^4,30,31^. Our analysis only included individuals of European ancestry with no melanoma history, similar to previously published nevus GWAS meta-analysis^4^.

### Polygenic risk score application cohorts

**KYAMS:** The KYAMS study was a follow-up of participants (aged 25-30 years) in the Kidskin Study, which began in 2015 to determine the impact of sun exposure intervention during childhood on myopia and eye health in adulthood^32^. The Kidskin study was initiated in 1995, a non-randomised controlled trial designed to reduce sun exposure in children through an educational intervention for children aged 5 to 6 years from 33 schools in Western Australia^33^. The KYAMS was approved by the Human Research Ethics Committees of the University of Western Australia (RA/4/1/6807) and written informed consent was obtained from all the participants^32^. For our analysis, we used the number of melanocytic nevi on the right arm counted by a trained observer. Biosamples for 113 participants were analysed in 2016, while 183 were analysed in 2021 using Infinium Global Screening Array. For quality control of the genotyped data, SNPs with a call rate <0.95, HWE p-value <10⁻⁶, and MAF (minor allele frequency) < 0.01 were excluded. Population outliers were identified using PCA (principal component analysis) based on participants of known European ancestry, using the 1000 Human Genome Project reference panel. Samples that were more than six standard deviations from the PC1 and PC2 centroids were excluded. One from each pair of individuals with an identity-by-descent score > 0.2 was removed. Post-quality control, additional genetic variants were imputed using the Trans-Omics for Precision Medicine (TOPMed) reference panel^34^. For our analysis, high-quality imputed data (R^2^ > 0.5 and MAF > 0.01) and nevus count data were available for 265 individuals.

**BTNS iris nevi:** The BTNS cohort (Supplementary table S1.1) has measurements for iris nevi in addition to the skin nevi used in the nevus GWAS meta-analysis. The cohort comprises a sample of predominantly adolescent twins (age 9-23 years) and their parents from Southeastern Queensland, Australia. The cohort was initially established to investigate the genetic factors influencing nevus development and other risk factors for melanoma^35,36^. The study was approved by the QIMR Human Research Ethics Committee and written informed consent was obtained from all participants. Genotyping was performed on Illumina 610-Quad arrays for 2,327 individuals phenotyped to the end of 2007 and on Illumina CoreExome arrays for 934 individuals phenotyped from 2008 to May 2013. After applying a series of quality controls, genotyped data were imputed using 1000 Genomes version 3 as described elsewhere^4^. The frequency of iris nevi derived from high-resolution digital photos of the iris was evaluated on an ordinal scale; (1) an absence of iris nevi, (2) at least one iris nevus, (3) at least three iris nevi, (4) at least five iris nevi, based on the number of melanin accumulations on the anterior border layer of the iris^37^. Although both the right and left iris were photographed, only the score from the right iris was included in the analysis due to the high polychoric correlation (0.91 to 0.95) between the two sides as reported in a previous study^37^. Post-quality-controlled (R^2^ > 0.5 and MAF > 0.01) imputed dosage data and iris nevus count data were available for 2,607 individuals.

### GWAS for QSkin I, QSkin II and AGDS

QSkin has two self-reported nevus measurements recorded as (1) a count on the left upper arm and (2) a 4-point categorical scale (“none”, “few”, “some”, “many”). The categorical nevus variable can be analysed as a linear or ordinal trait. We used QSkin I data to compare the performance of a linear model in Scalable and Accurate Implementation of the GEneralised mixed model (SAIGE)^38^ and the proportional odds logistic mixed model (POLMM) which is implemented for ordinal trait analysis^39^. Both methods control sample relatedness. The performance of these GWAS models was assessed by comparing the number of significant hits and the total SNP heritability estimated using linkage disequilibrium score regression (LDSC)^40^. A linear model in SAIGE was the best-performing method (Supplementary Notes S1.2 and Supplementary Figure 1-2) and was then used to conduct GWAS in the QSkin II (Supplementary Figure 4) and AGDS cohorts (Supplementary Figure 5).

### Heritability and LD score intercept estimation

We estimated the proportion of variance explained by all SNPs (SNP-based heritability) for nevus count using QSkin I through genome-based restricted maximum likelihood (REML) implemented in GCTA v.1.26^41^. Following quality control, a genetic relationship matrix (GRM) was constructed using genotyped SNPs with MAF > 0.01. The GRM was fitted in a REML analysis with 10 PCs included as covariates; a GRM cutoff of 0.05 was then applied to exclude related people.

LDSC regression (v1.0.0) was conducted for nevus GWAS summary results using pre-calculated LD scores from a 1000 Genomes European reference population, to estimate the genomic inflation factor, and to estimate the SNP-based heritability for GWAS where only summary statistics were available^40^.

### GWAS meta-analysis

Our meta-analysis was based on 14 GWASs of nevus count. As discussed in the cohort details (Supplementary Notes S1.2), there were slight differences in the assessment of nevus count, individual-level genotyping, imputation and quality control criteria, and the statistical models and analytical software employed in each GWAS. Therefore, we conducted a preliminary analysis to demonstrate that each study was consistent for conducting a GWAS meta-analysis for the nevus count trait. Specifically, after aligning effect alleles in all studies, we performed inverse variance weighted Mendelian Randomisation (IVW MR)^42^ as implemented in the “MendelianRandomisation” R package. The nevus count of each QSkin I, QSkin II and AGDS study was obtained on a similar 4-point scale self-reported phenotypic assessment with moderate to high levels of reproducibility^43,44^ and the effect sizes of genome-wide significant SNPs were consistent across these three GWASs (Supplementary Figure 18). Hence, we used the GWAS meta-analysis conducted for QSkin I, QSkin II and AGDS (Supplementary Figure 6) as the reference study, and its top independent genome-wide significant SNPs as the instrumental variables to perform IVW MR. Based on the study-specific IVW MR plots (Supplementary Figure 7-17), all individual studies from Duffy et al. (2018) were consistent with the three new studies.

Because the measurement of nevus count varies across studies, the meta-analysis of nevus GWAS was conducted using a fixed-effect sample size weighted Z-score method as implemented in METAL v.2020-05-05^45^. We applied genomic control correction to input GWAS summary statistics. For post-GWAS analysis that required effect sizes and standard errors, for each SNP, we estimated the effect size and standard error using the Z-statistics as these were the only outputs available^46^.

Before conducting any post-GWAS analyses, the following quality control steps were applied to the meta-GWAS summary statistics. First, GWAS variants driven by a single study were dropped to ensure SNPs were associated in multiple studies. The DENTIST (Detecting Errors iN analyses of summary staTISTtistics) method (v.1.3.0.0), was used with default settings to obtain improved GWAS summary statistics by reducing heterogeneity between GWAS summary data and LD reference data^47^. In DENTIST analysis, LD structure was obtained using a UK Biobank European reference panel comprising 5,000 randomly selected and unrelated individuals. In this step, 8,783,913 variants were tested by applying the DENTIST method and 124,531 variants were filtered out.

Then, the FUMA (Functional Mapping and Annotation of GWASs) platform v.1.6.0^48^ was used to identify independent significant SNPs and significant loci using pre-calculated LD structure from the 1000 Genomes Phase 3 European reference population *(n* = 503). Independently significant SNPs were defined as genome-wide significant (< 5×10^−8^) SNPs not in LD at R^2^ < 0.1. Independent lead SNPs were characterised as independently significant SNPs not in LD at R^2^ < 0.05. Genomic loci associated with nevus count are defined by merging lead SNPs within a 1 Mb window, along with all SNPs with LD of R^2^ ≥ 0.05 with any independent SNPs. In addition, candidate SNPs that were defined as the SNPs having R^2^ ≥ 0.1 with one of the independent significant SNPs and MAF ≥ 0.01, were used for further annotations in FUMA. Forest plots of genome-wide significant lead SNPs were observed to check that the effect sizes were consistent across studies (Supplementary Figure 18).

### Functional annotation, gene-based test and gene-set analysis

Candidate SNPs were annotated for functional consequences on genes based on Ensembl genes [build 85] annotated using ANNOVAR in FUM ^48^.

Then positional, expression quantitative trait loci (eQTL) and chromatin interaction mapping approaches were conducted in FUMA to prioritise genes. First, annotated SNPs were mapped to genes considering the physical position of the SNP inside a gene using a 10 kb window. Further, eQTL mapping was conducted within a 1 Mb window (cis-eQTL) to test whether GWAS SNPs are associated with the expression of the gene at a false discovery rate (FDR) ≤ 0. 05. eQTL data included were restricted to skin tissue from TwinsUK and Genotype-Tissues Expression (GTEx) v8 (sun-exposed, not sun-exposed skin types and fibroblasts). Moreover, chromatin interaction mapping was performed to test whether there was a significant (FDR ≤ 1×10^−6^) chromatin interaction between nevus-associated regions and the promoter of nearby genes. Promoters were defined as 250 bp upstream and 500 bp downstream of the transcription site and data were included from 14 tissues (e.g. ovary, bladder, aorta and lung) and seven cell types (e.g. embryonic stem cell, liver, mesenchymal stem, and fibroblast).

In addition, MAGMA (Multi-marker Analysis of GenoMic Annotation)^49^ gene-based and gene-set analyses were carried out as implemented in FUMA^48^. In the gene-based analyses, SNPs were mapped to 18,915 protein-coding genes obtained from Ensembl build 85. After accounting for LD, the association test statistics for SNPs were combined into a single gene-level p-value. Gene-wide significance was defined at P = 0.05/18915 = 2.64×10^−6^.

Here we conducted two separate gene-set analyses using candidate genes selected from MAGMA gene-based analysis and three gene mapping approaches. In the MAGMA gene-set analysis, MAGMA gene-based p-values were further combined into each gene set from 4,761 curated gene sets and 5,917 gene ontology (go) terms from MsigDB v2023^48^. Significant gene sets were selected based on Bonferroni-corrected P-values. In addition to MAGMA gene-set analysis, genes prioritised by any of three gene mapping techniques were tested for overrepresentation in gene sets obtained from MsigDB, WikiPathways and the GWAS catalog if at least two prioritised genes overlapped with a testing gene set^48^. P-values of gene sets were adjusted using the Benjamini-Hochberg correction.

### Transcriptome-wide association study and colocalisation analysis for nevus count

As an alternative approach to identifying candidate genes, a TWAS was performed for the nevus GWAS meta-analysis. eQTL data was sourced from skin tissue in GTex v8 (sun-exposed skin; *n* = 508, not-sun-exposed skin; *n* = 430, and fibroblasts; *n* = 403), whole-blood (*n* = 31,684) from eQTLGen^50^ and a set of 106 primary melanocyte samples^5,51^. For melanocytes and other skin tissues in GTEx, pre-computed gene expression weights are available in a previous study^5,51^ and FUSION website (http://gusevlab.org/projects/fusion/) respectively, and we performed TWAS for these tissues in FUSION using them. For whole-blood tissue, only eQTL summary-level data are available from the eQTLGen consortium. Since FUSION requires individual-level expression data to compute expression weights, we performed TWAS for whole-blood tissue in SUMMIT v.1.0.2 (Summary-level Unified Method for Modeling Integrated Transcriptome) which supports summary-level expression data^52^. This analysis utilised pre-calculated gene expression imputation models in SUMMIT specifically built for eQTLGen data (https://doi.org/10.17605/OSF.IO/7MXSA).

For each set, a Bonferroni corrected p-value threshold was defined as 0.05/(no of tested genes) in all five tissue sets (melanocytes; 3797genes, three skin tissues; (9573+9556+10661) = genes, whole blood; 11415 genes, P-value is 0.05/(3797+9573+9556+10661+11415) = 1.11e-6).

Colocalisation analysis was conducted to estimate the posterior probability of a shared putative causal genetic variant between gene expression and nevus count (if PP4 ≥ 0.75), as a complementary analysis for TWAS ^53^. For melanocytes and skin tissues, colocalisation analyses were conducted within FUSION which includes an interface to COLOC v.5.1.0.1 R package^54^. A p-value threshold (--coloc_P flag) of 0.05 was used to compute COLOC statistics in FUSION. For eQTLGen data, colocalisation analysis was performed using COLOC v.5.1.0.1 package in R v.4.2.0.

### GWAS-PW for nevus count and melanoma

GWAS-PW v.0.21^9^ was used on nevus GWAS meta-analysis data and previously published melanoma GWAS^5^ to determine whether the significant genetic loci from nevus GWAS meta-analysis were pleiotropic for nevus count and melanoma. For both nevus count and melanoma, SNPs were assigned to LD blocks using the recommended boundaries provided by GWAS-PW (https://bitbucket.org/nygcresearch/ldetect-data). GWAS-PW estimates posterior probability (PPA) for four different models: a locus (i) has an association with nevus count only; (ii) has an association with melanoma only; (iii) has a shared association with both melanoma and nevus count through the same genetic variant; (iv) is associated with both melanoma and nevus count through independent genetic variants. Any genome-wide significant locus with PPA > 0.8 for hypothesis (iii) was defined as a pleiotropic locus for melanoma and nevus count.

### Polygenic risk score for nevus count

A polygenic risk score (PRS) for nevus count was derived using SBayesRC which uses functional genomic annotations to improve polygenic prediction^55^. To generate PRS for testing in adults and adolescents, as a discovery dataset, we conducted a nevus GWAS meta-analysis using 13 out of 14 cohorts excluding QSkin II (final *n* = 79,357), whereas to predict iris nevus count, a separate (cutaneous) nevus GWAS meta-analysis was conducted excluding BTNS (final *n* = 82,704) as above. In addition, LD data calculated from UK Biobank European ancestry and functional annotation data provided by SBayesRC (https://github.com/zhilizheng/SBayesRC) were used in SBayesRC analysis.

We used QSkin II (*n* = 6,608), KYAMS (*n* = 265) and BTNS (*n* = 2,607) cohorts to determine the ability of nevus PRS to predict cutaneous nevus count in older adults (average age = 63 years, SD = 13 years), young adults (average age = 28 years, SD = 1 year), and then iris nevus count respectively. To score individuals in independent cohorts, imputed allelic dosages were weighted by the SBayesRC-derived SNP weights using PLINK v.2.0 (www.cog-genomics.org/plink/2.0/). We then standardised the nevus PRS to have a mean of zero and SD of 1. Since related individuals were retained in all cohorts, GCTA GREML was then used to regress inverse-normal rank transformed cutaneous nevus count/iris nevus count, adjusted for sex, age, age^2^, sex*age, sex*age^2^, the first 10 PCs and a GRM, onto standardised PRS. For Kidskin and BTNS cohorts, we used a binary batch variable as an additional covariate, to control the genotype batch effect. Finally, nevus PRS performance was determined using R^2^ computed applying a formula reported in^56^.

## Acknowledgments

S.M. is supported by an Investigator grant (2034568) from the Australian National Health and Medical Research Council (NHMRC). M.M.I is supported in part by the National Institute for Health and Care Research (NIHR) Leeds Biomedical Research Centre (BRC) (NIHR203331). The views expressed are those of the author(s) and not necessarily those of the NHS, the NIHR or the Department of Health and Social Care. S.E.M. is supported by NHMRC grants APP1172917 and APP2025674. N.K.H. is supported by the NHMRC (APP1195581), Jan Brown, the Buck Off Melanoma community and memorial donors honouring Nicola Laws. This work was conducted using the UK Biobank Resource (application number 25331). We acknowledge the contribution of the data from the published nevus meta-analysis in 2018^4^ and the melanoma meta-analysis in 2020^5^. All studies are forever grateful for the invaluable contributions of all research participants, their families, research nurses, research assistants, and support staff, whose dedication made this work possible. Please see supplementary notes for more details of acknowledgments and financial support for contributing studies.

## Author contribution

M.H.L conceived the study and designed the analysis. M.H.L. and S.M. jointly supervised the study. G.J.M.S.R.J. carried out the analysis. G.J.M.S.R.J. and M.H.L. wrote the first draft of the manuscript. G.Z., N.P., C.M.O., N.G.M., P.A.L., S.E.M., S.D.G., S.D.T., G.L., S.S.Y.L., T.N., M.K., L.M. P., G.W.M., N.K.H., J.M.P., D.J.H., J.H., A.W.H., M.F., D.T.B., K.M.B., V.B., D.A.M., M.M.I., D.C.W., D.L.D. were involved in cohort data collection. All authors contributed to the final version of the manuscript.

## Data availability

The summary data for the GWAS meta-analysis generated in this study is available on Zenodo [URL to be confirmed]. Individual-level data from UK Biobank was accessed under application number 25331. Access to UK Biobank data can be obtained by applying to UK Biobank (https://www.ukbiobank.ac.uk/). All other data supporting this study may be available to the corresponding authors upon request, subject to ethical and regulatory approval from the relevant institution.

## Code availability

The codes used in this study are available upon request to the corresponding author

