## Supplementary Figures and Notes for "A large-scale genome-wide association meta-analysis for nevus count provides direct insights into the genetics of melanoma"

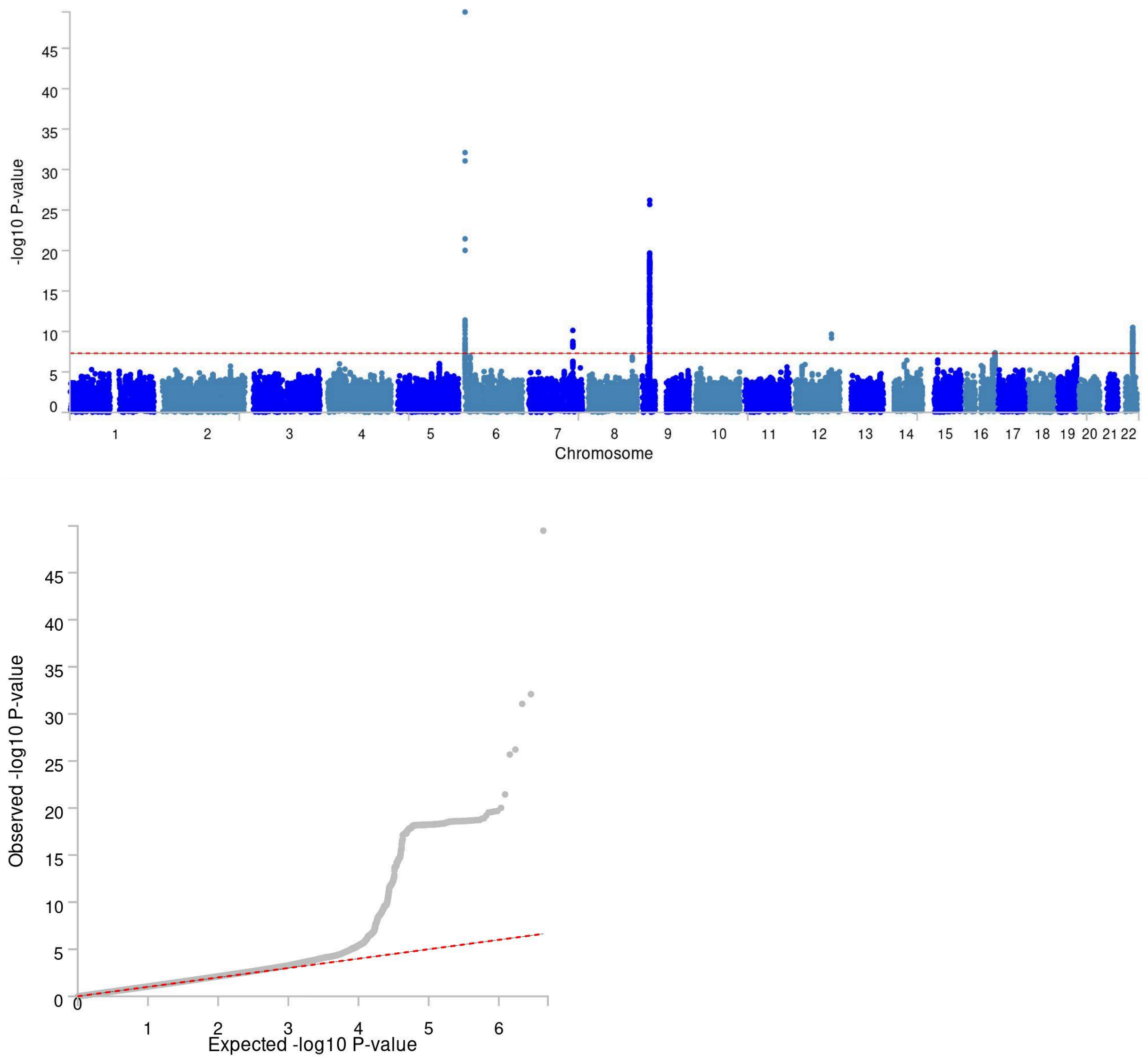


Supplementary Figure 1: Manhattan plot and Quantile-Quantile plot of the GWAS of 4-point scale nevus count from the QSkin I obtained applying SAIGE method. ($n$ = 15,346). The genomic inflation factor was 1.06. SNP-based heritability estimated by LDSC regression was 0.16 (0.037).

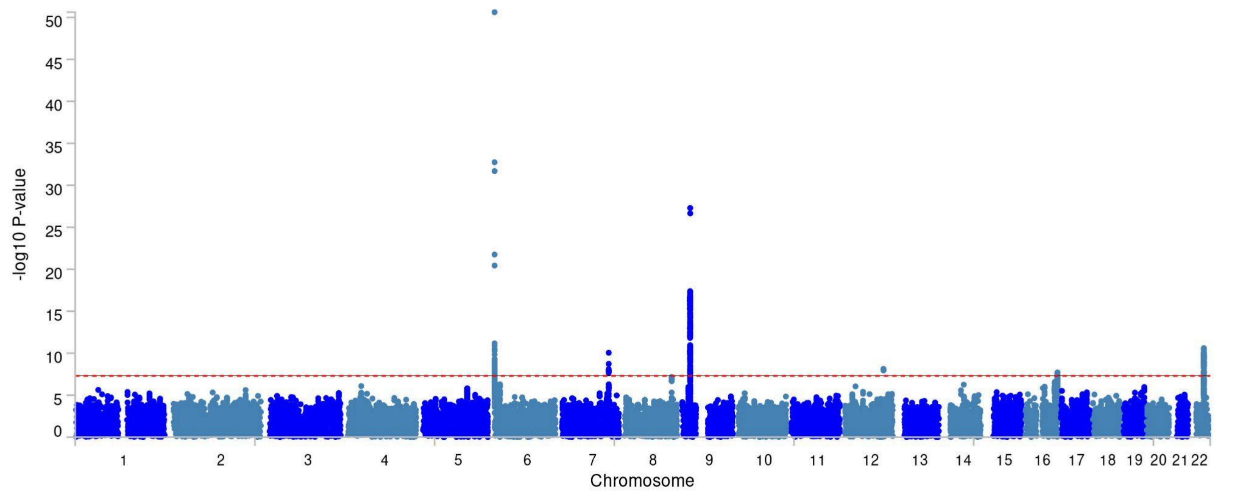


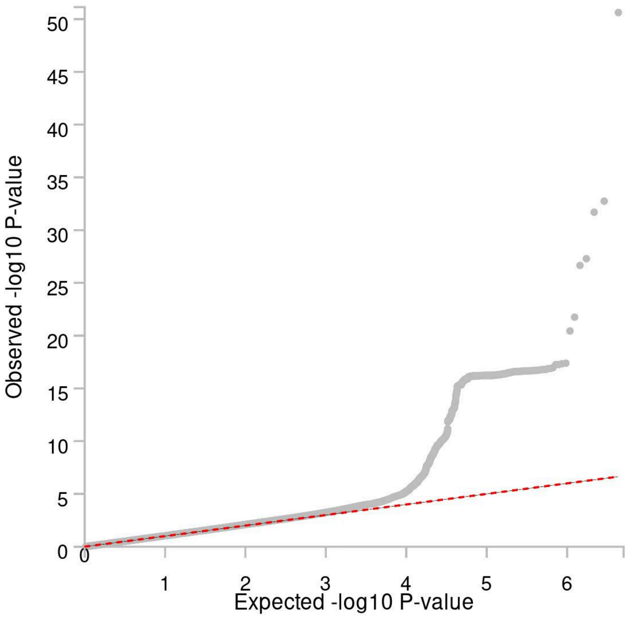


Supplementary Figure 2: Manhattan plot and Quantile-Quantile plot of the GWAS of 4-point scale nevus count from the QSkin I obtained applying POLMM method. (*n* = 15,346). The genomic inflation factor was 1.05. SNP-based heritability estimated by LDSC regression was 0.13 (0.036).

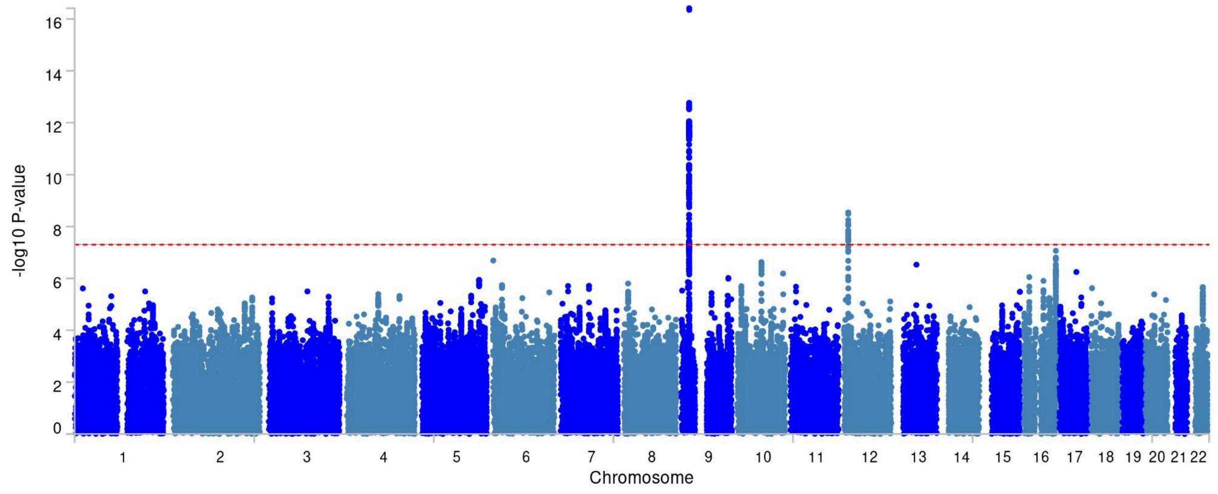


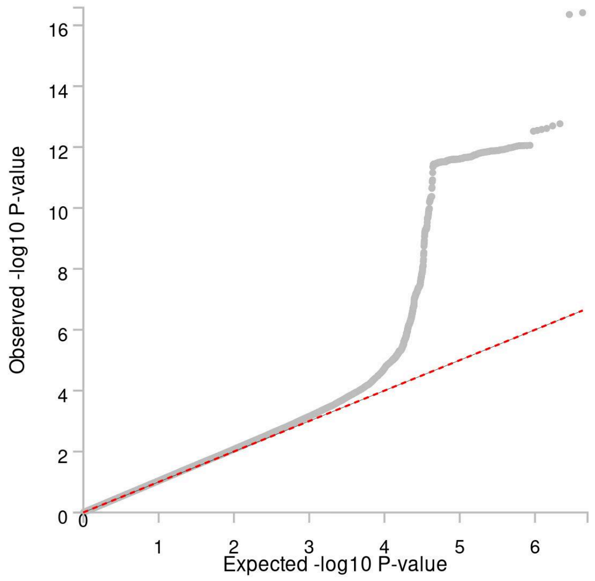


Supplementary Figure 3: Manhattan plot and Quantile-Quantile plot of the GWAS of quantitative nevus count from the QSkin I obtained applying SAIGE method. (*n* = 13,007). The genomic inflation factor was 1.07. SNP-based heritability estimated by LDSC regression was 0.18 (0.045).

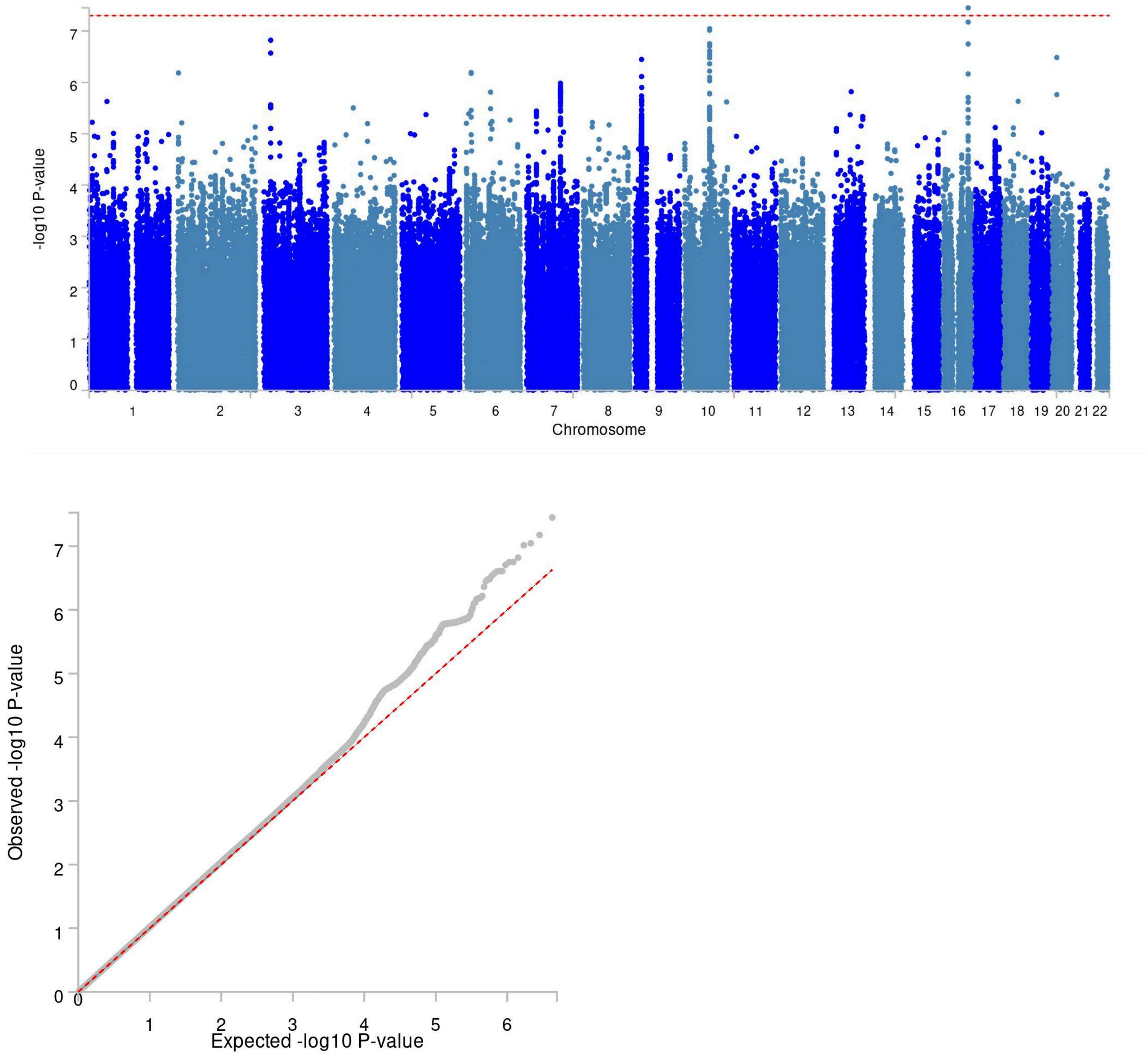


Supplementary Figure 4: Manhattan plot and Quantile-Quantile plot of the GWAS of the 4-point scale nevus count from QSkin II applying SAIGE method. ($n$ = 6,608). The genomic inflation factor was 1.04. SNP-based heritability estimated by LDSC regression was 0.03 (0.067).

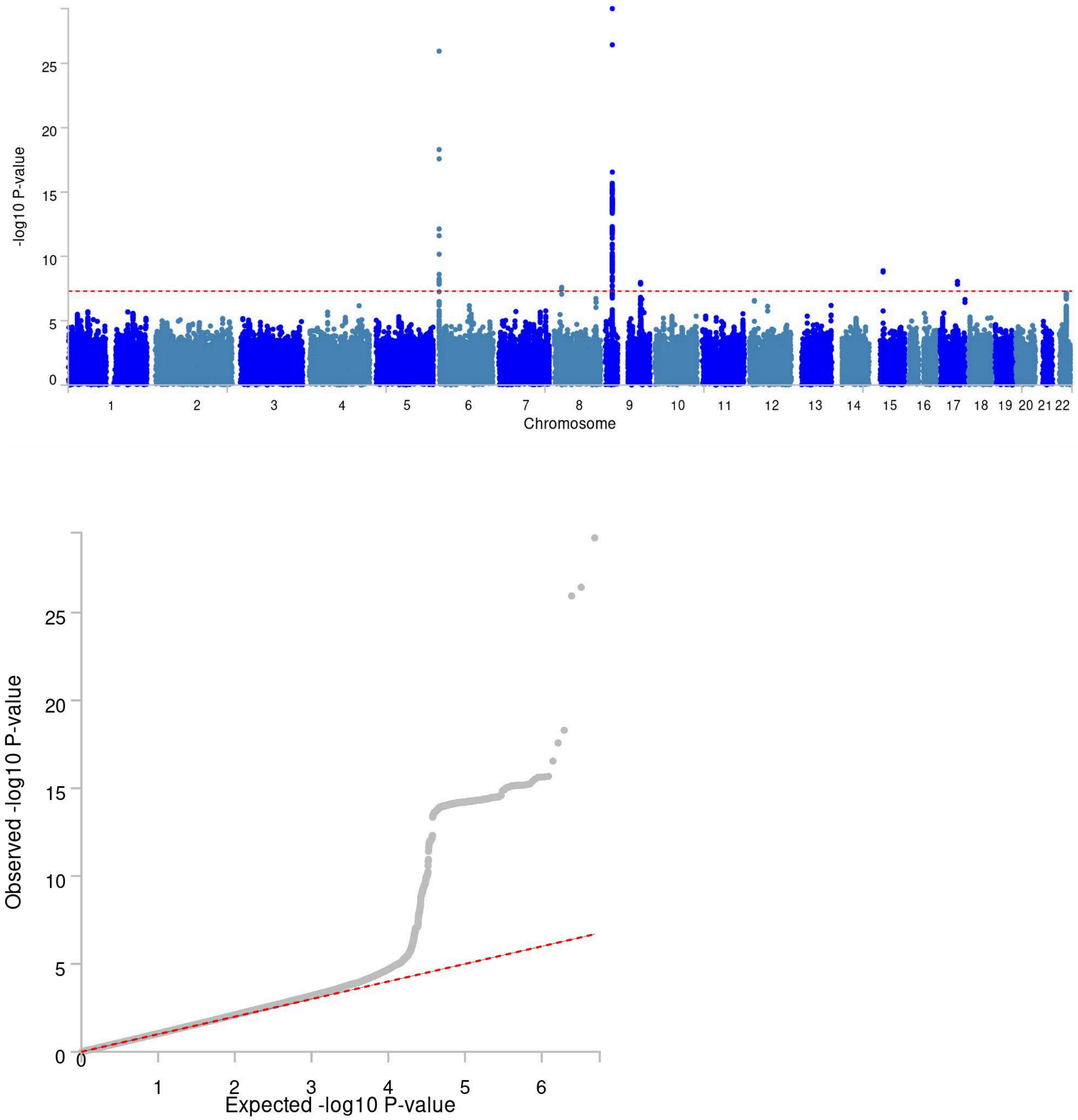


Supplementary Figure 5: Manhattan plot and Quantile-Quantile plot of the GWAS of the 4-point scale nevus count from AGDS applying SAIGE method. (*n* = 11,775). The genomic inflation factor was 1.06. SNP-based heritability estimated by LDSC regression was 0.11 (0.042).

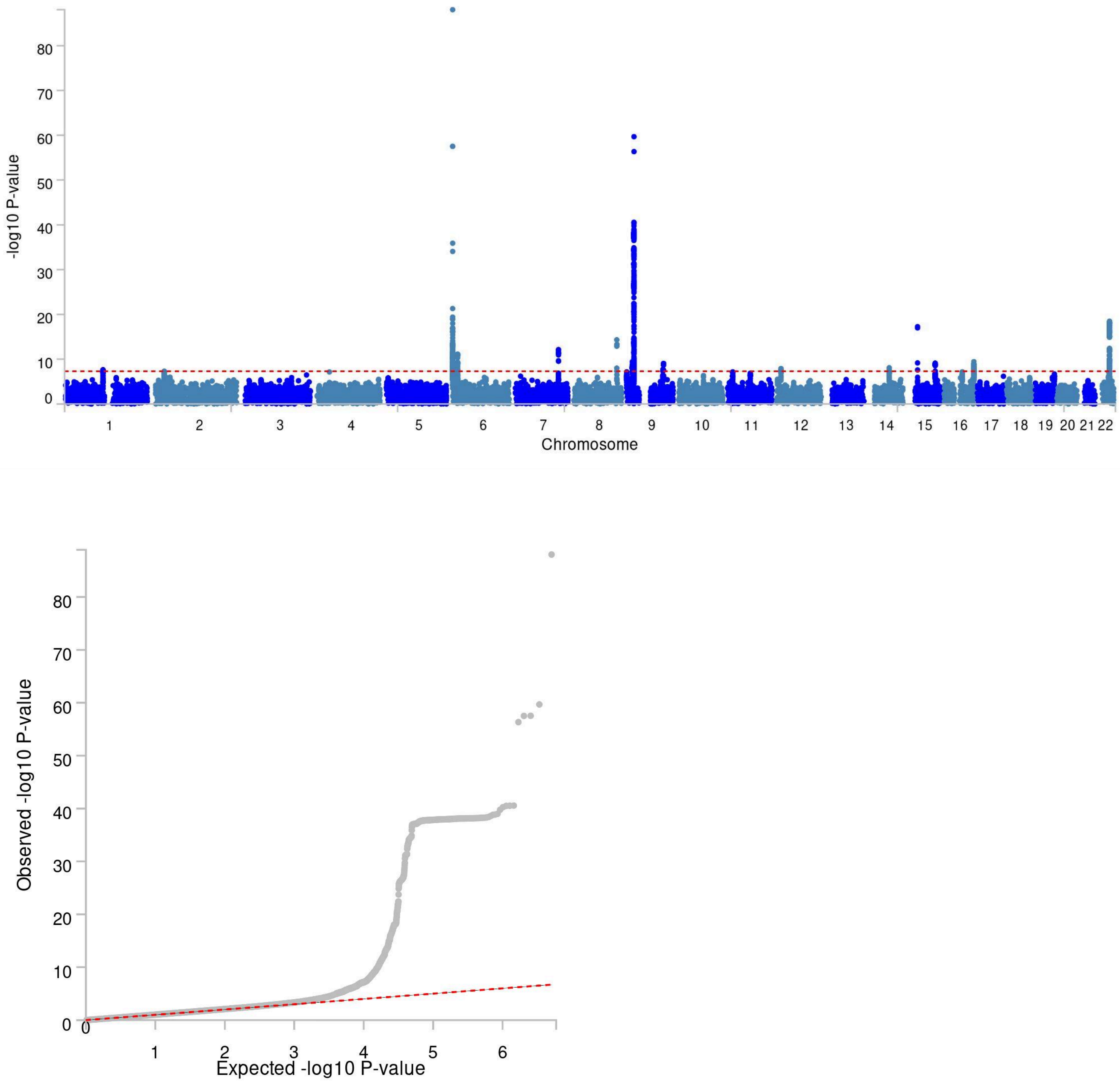


Supplementary Figure 6: Manhattan plot and Quantile-Quantile plot of the GWAS meta-analysis of the 4-point scale nevus count from QSkin I, QSkin II and AGDS applying fixed-effect inverse variance weighted method. (*n* = 33,729). The genomic inflation factor was 1.06. Total SNP-heritability estimated from LDSC regression was 0.12 (0.02)

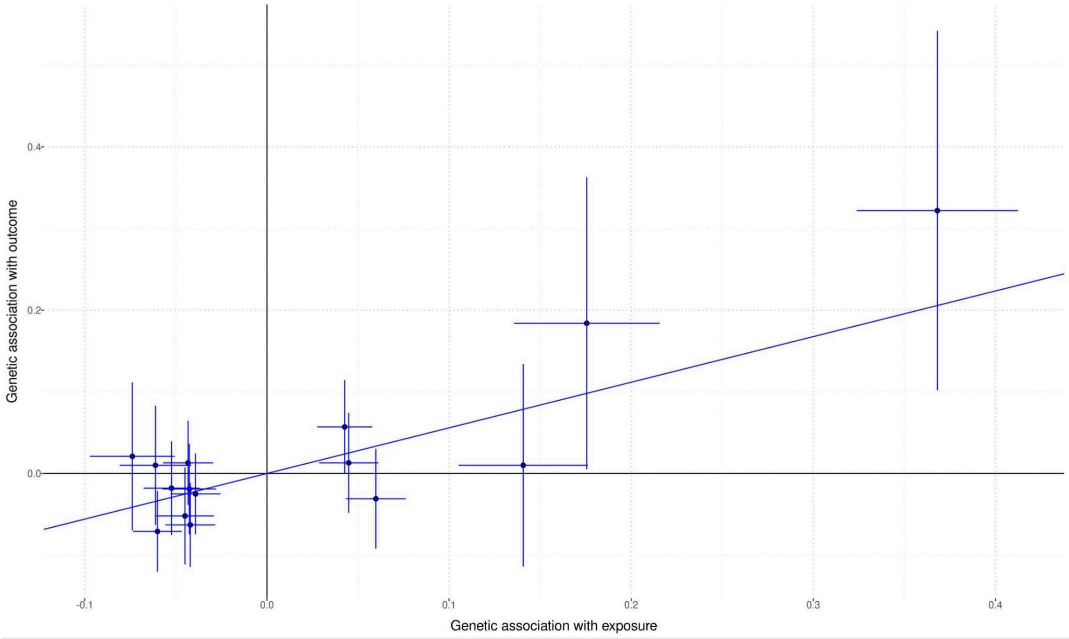


Supplementary Figure 7: IVW MR plot for ALSPAC study. Exposure is GWAS meta-analysis summary results of QSkin I, QSkin II and AGDS and the outcome is GWAS summary results from the ALSPAC study. Instrumental variables were significant leading hits from GWAS meta-analysis of QSkin I, QSkin II and AGDS.

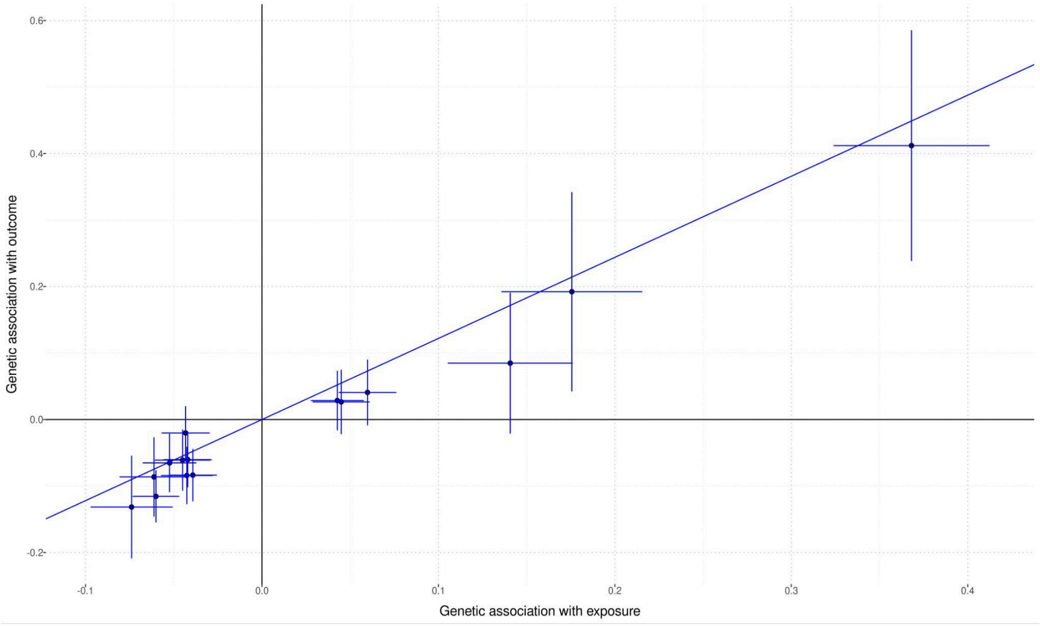


Supplementary Figure 8: IVW MR plot for Harvard study. Exposure is GWAS meta-analysis summary results of QSkin I, QSkin II and AGDS and outcome is GWAS summary results from Harvard study.

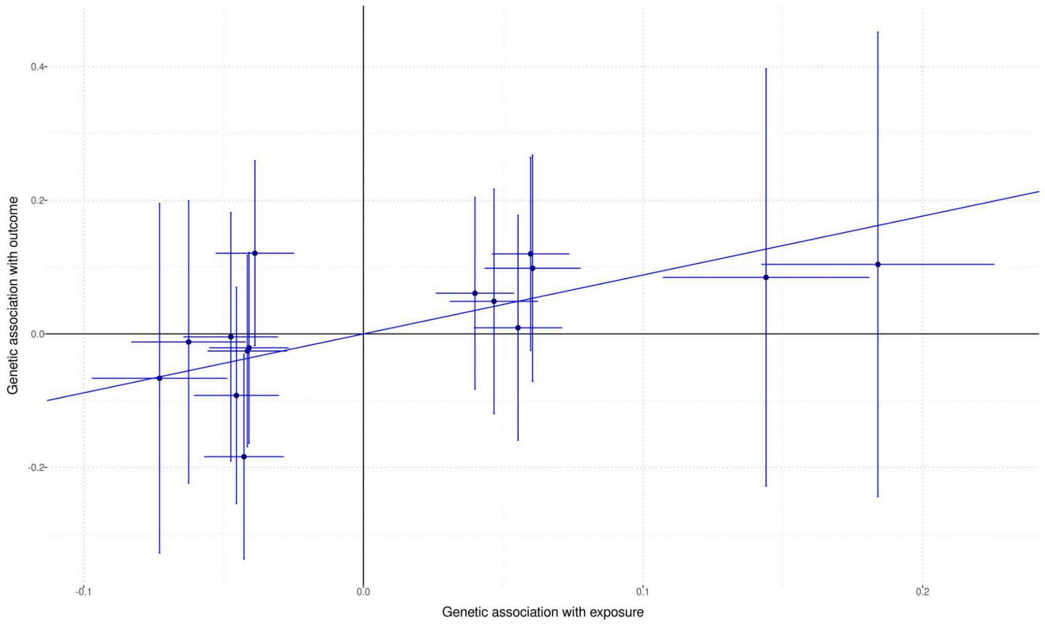


Supplementary Figure 9: IVW MR plot for Leeds study. Exposure is GWAS meta-analysis summary results of QSkin I, QSkin II and AGDS and the outcome is GWAS summary results from Leeds study.

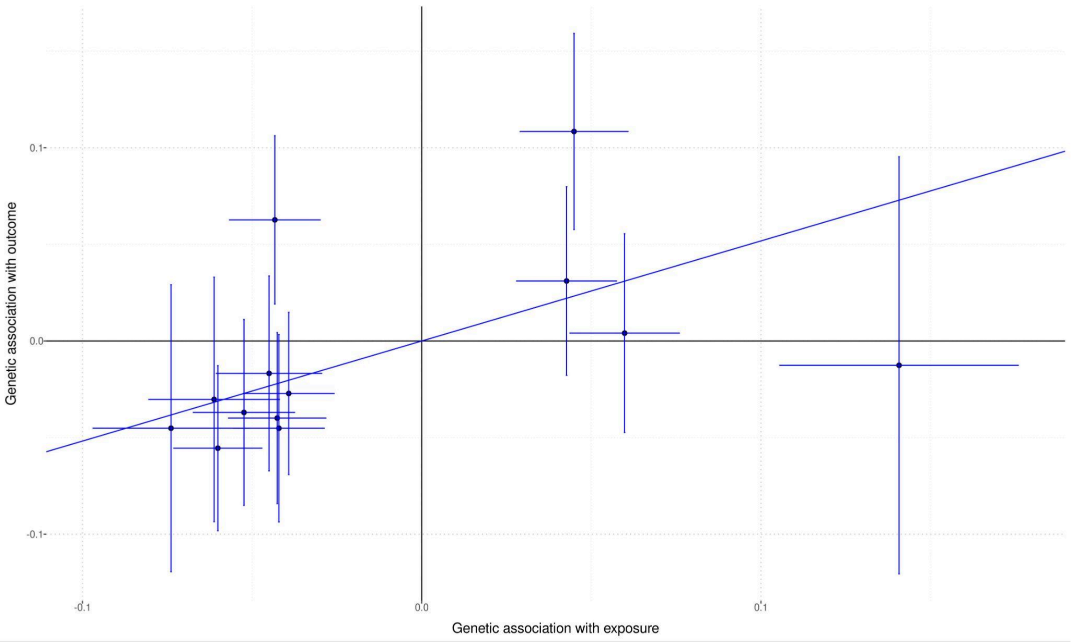


Supplementary Figure 10: IVW MR plot for BTNS study. Exposure is GWAS meta analysis summary results of QSkin I, QSkin II and AGDS and outcome is GWAS summary results from BTNS study.

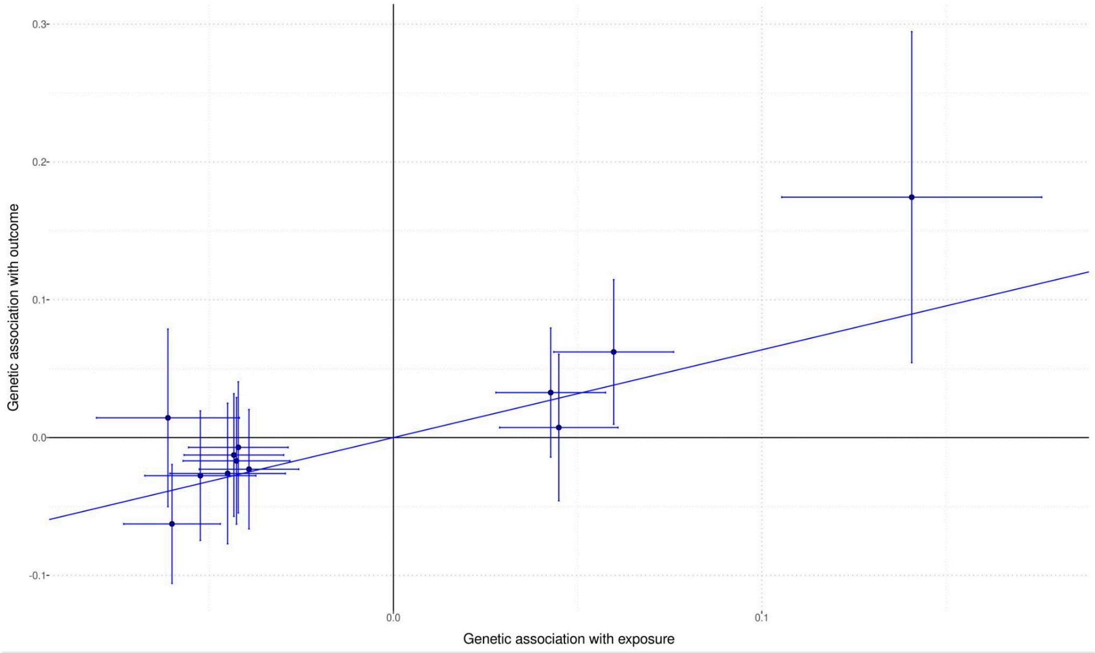


Supplementary Figure 11: IVW MR plot for Parents of BTNS Twins study. Exposure is GWAS meta analysis summary results of QSkin I, QSkin II and AGDS and outcome is GWAS summary results from Parents of BTNS Twins study.

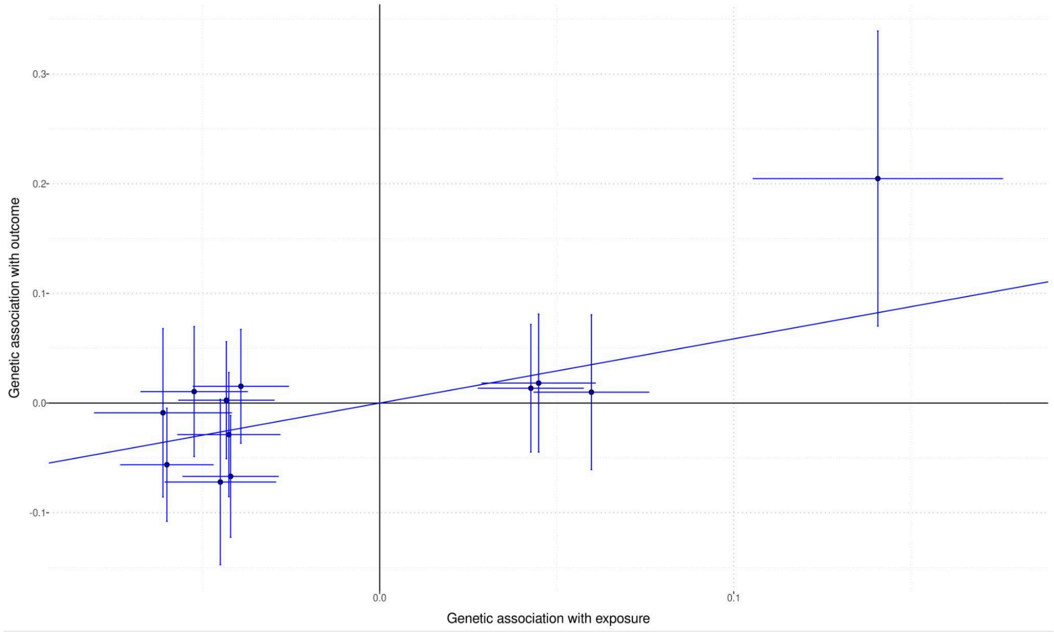


Supplementary Figure 12: IVW MR plot for QIMR Adult Twins study. Exposure is GWAS meta analysis summary results of QSkin I, QSkin II and AGDS and outcome is GWAS summary results from QIMR Adult Twins study.

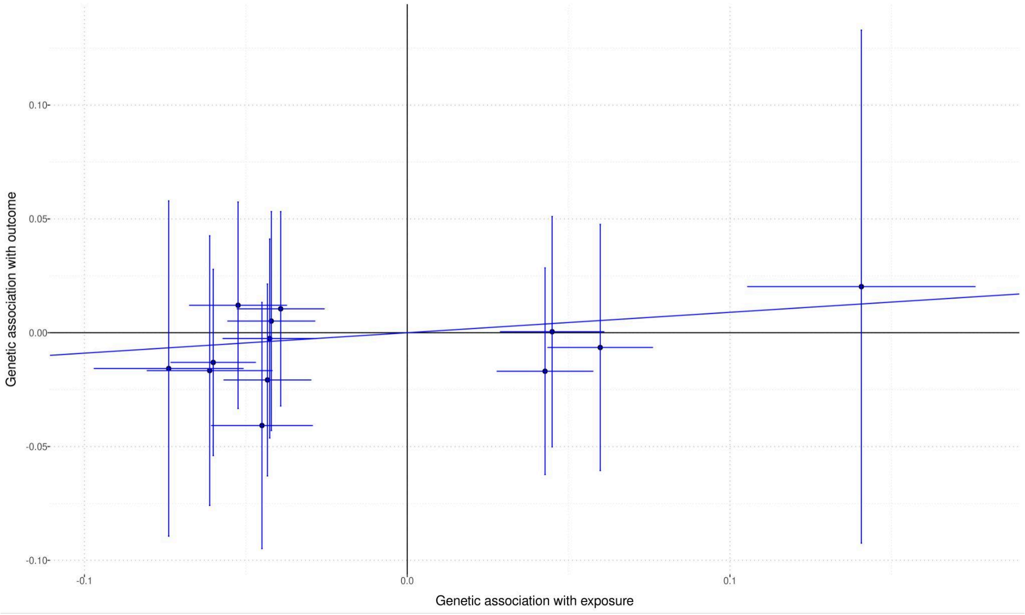


Supplementary Figure 13: IVW MR plot for QIMR Twins > 50 years study. Exposure is GWAS meta-analysis summary results of QSkin I, QSkin II and AGDS and outcome is GWAS summary results from QIMR Twins > 50 years study.

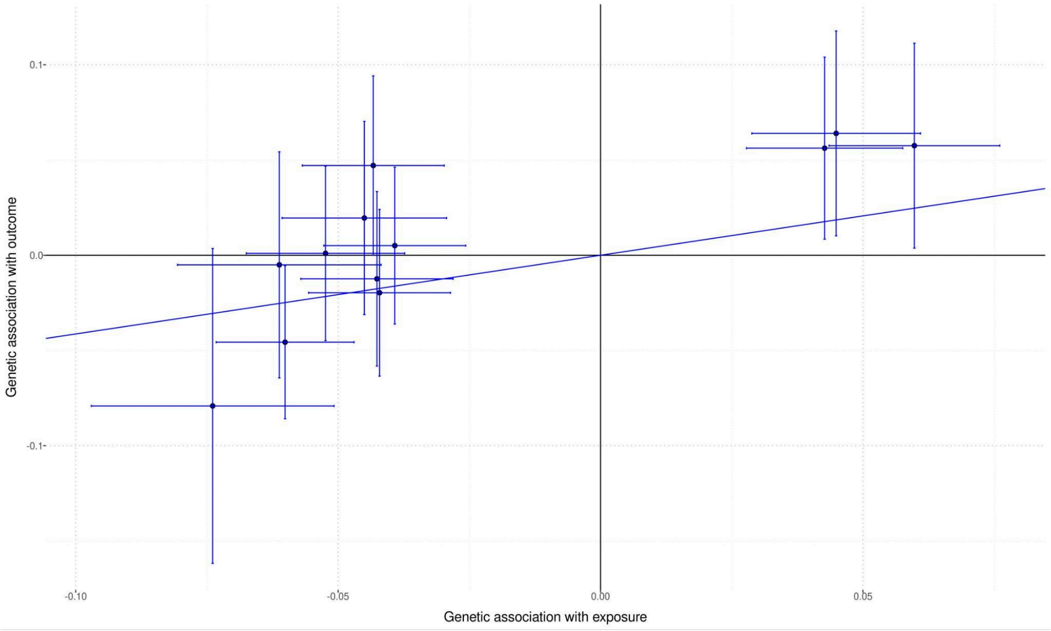


Supplementary Figure 14: IVW MR plot for the Raine Study. Exposure is GWAS meta-analysis summary results of QSkin I, QSkin II and AGDS and outcome is GWAS summary results from the Raine Study.

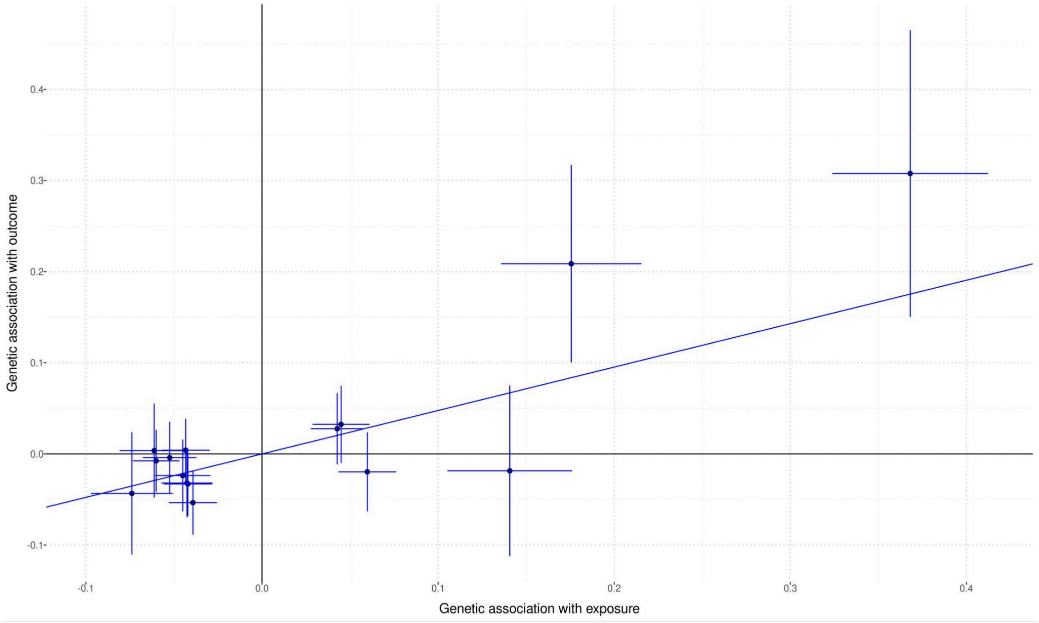


Supplementary Figure 16: IVW MR plot for Rotterdam Study. Exposure is GWAS meta-analysis summary results of QSkin I, QSkin II and AGDS and the outcome is GWAS summary results from Rotterdam Study.

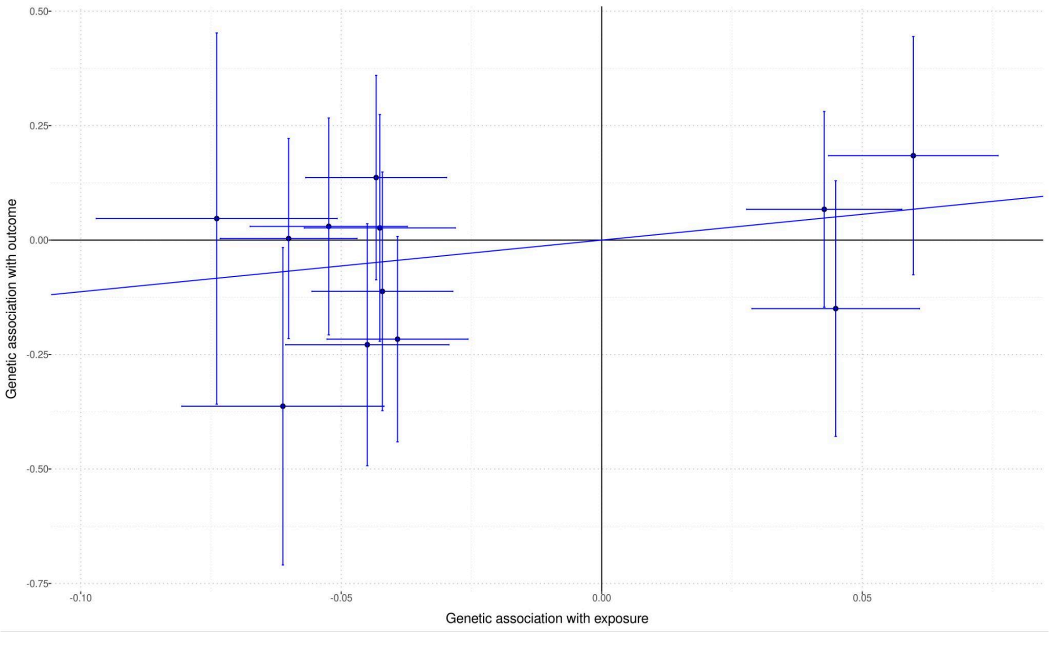


Supplementary Figure 15: IVW MR plot for Twins Eyes Study in Tasmania Study. Exposure is GWAS meta-analysis summary results of QSkin I, QSkin II and AGDS and outcome is GWAS summary results from Twins Eyes Study in Tasmania Study.

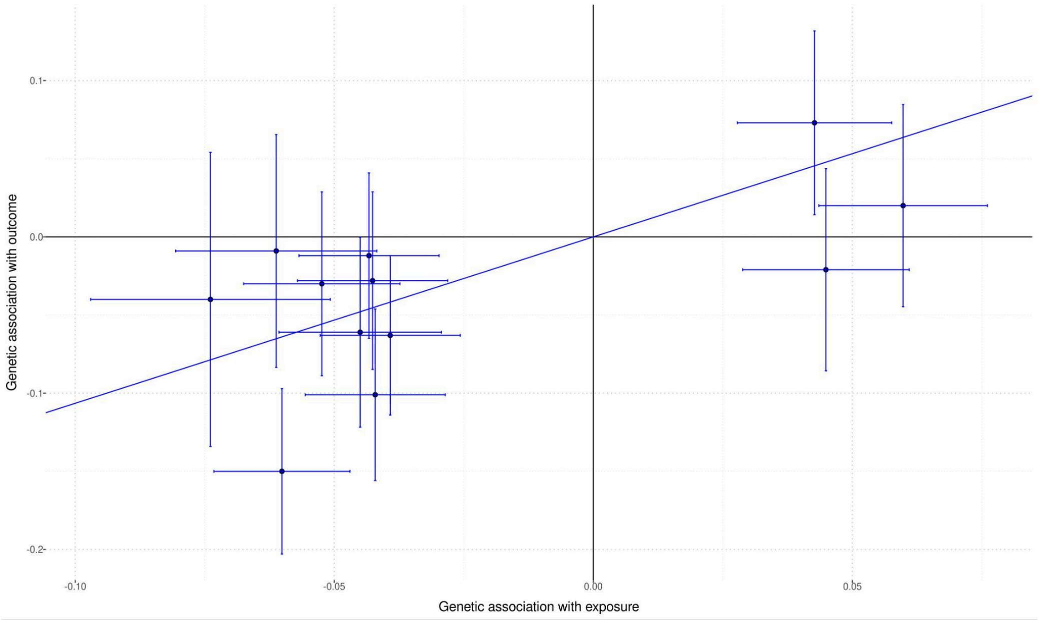


Supplementary Figure 17: IVW MR plot for TwinsUK Study. Exposure is GWAS meta-analysis summary results of QSkin I, QSkin II and AGDS and the outcome is GWAS summary results from TwinsUK Study.

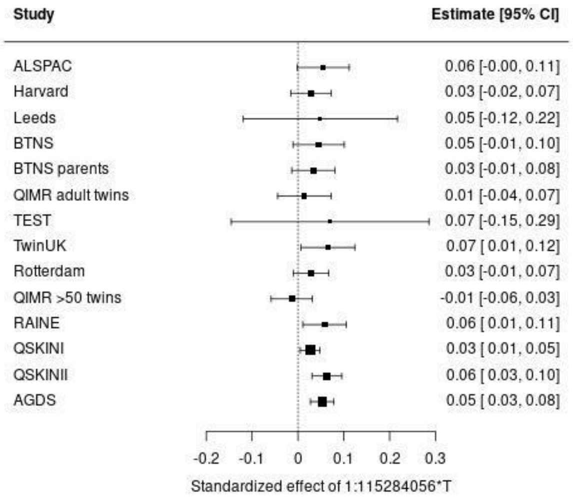
Study
Estimate [95% CI]
ALSPAC 0.06 [-0.00, 0.11]
Harvard 0.03 [-0.02, 0.07]
Leeds 0.05 [-0.12, 0.22]
BTNS 0.05 [-0.01, 0.10]
BTNS parents 0.03 [-0.01, 0.08]
QIMR adult twins 0.01 [-0.04, 0.07]
TEST 0.07 [-0.15, 0.29]
TwinUK 0.07 [0.01, 0.12]
Rotterdam 0.03 [-0.01, 0.07]
QIMR >50 twins -0.01 [-0.06, 0.03]
RAINE 0.06 [0.01, 0.11]
QSKINI 0.03 [0.01, 0.05]
QSKINII 0.06 [0.03, 0.10]
AGDS 0.05 [0.03, 0.08]
-0.2 -0.1 0 0.1 0.2 0.3
Standardized effect of 1:115284056*T

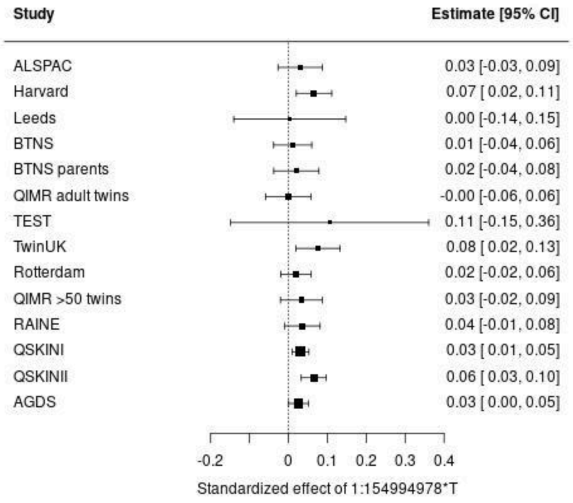
Study
Estimate [95% CI]
ALSPAC 0.03 [-0.03, 0.09]
Harvard 0.07 [0.02, 0.11]
Leeds 0.00 [-0.14, 0.15]
BTNS 0.01 [-0.04, 0.06]
BTNS parents 0.02 [-0.04, 0.08]
QIMR adult twins -0.00 [-0.06, 0.06]
TEST 0.11 [-0.15, 0.36]
TwinUK 0.08 [0.02, 0.13]
Rotterdam 0.02 [-0.02, 0.06]
QIMR >50 twins 0.03 [-0.02, 0.09]
RAINE 0.04 [-0.01, 0.08]
QSKINI 0.03 [0.01, 0.05]
QSKINII 0.06 [0.03, 0.10]
AGDS 0.03 [0.00, 0.05]
-0.2 0 0.1 0.2 0.3 0.4
Standardized effect of 1:154994978*T

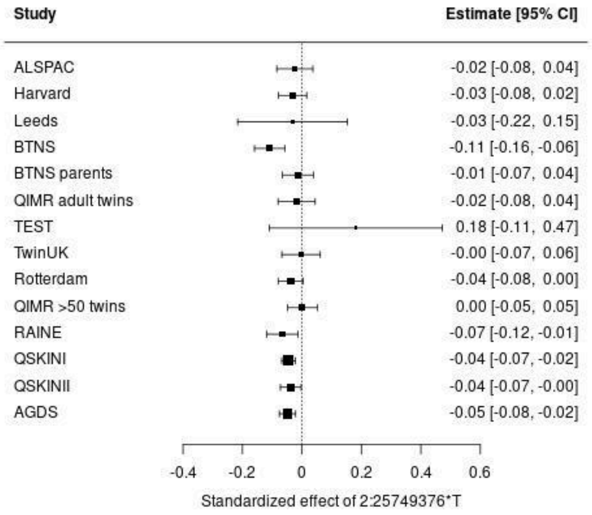
Study
Estimate [95% CI]
ALSPAC -0.02 [-0.08, 0.04]
Harvard -0.03 [-0.08, 0.02]
Leeds -0.03 [-0.22, 0.15]
BTNS -0.11 [-0.16, -0.06]
BTNS parents -0.01 [-0.07, 0.04]
QIMR adult twins -0.02 [-0.08, 0.04]
TEST 0.18 [-0.11, 0.47]
TwinUK -0.00 [-0.07, 0.06]
Rotterdam -0.04 [-0.08, 0.00]
QIMR >50 twins 0.00 [-0.05, 0.05]
RAINE -0.07 [-0.12, -0.01]
QSKINI -0.04 [-0.07, -0.02]
QSKINII -0.04 [-0.07, -0.00]
AGDS -0.05 [-0.08, -0.02]
-0.4 -0.2 0 0.2 0.4 0.6
Standardized effect of 2:25749376*T

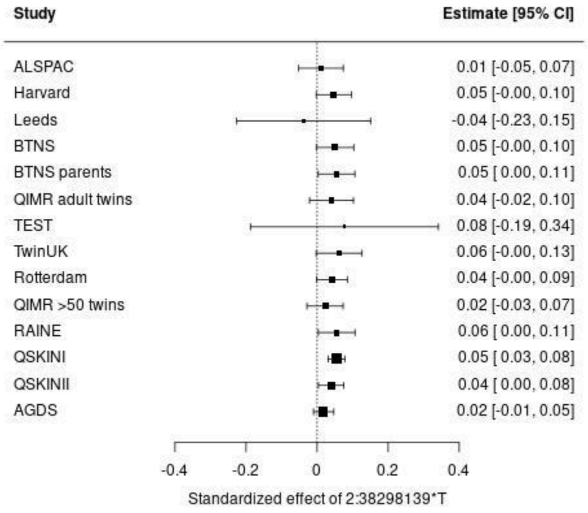
Study
Estimate [95% CI]
ALSPAC 0.01 [-0.05, 0.07]
Harvard 0.05 [-0.00, 0.10]
Leeds -0.04 [-0.23, 0.15]
BTNS 0.05 [-0.00, 0.10]
BTNS parents 0.05 [0.00, 0.11]
QIMR adult twins 0.04 [-0.02, 0.10]
TEST 0.08 [-0.19, 0.34]
TwinUK 0.06 [-0.00, 0.13]
Rotterdam 0.04 [-0.00, 0.09]
QIMR >50 twins 0.02 [-0.03, 0.07]
RAINE 0.06 [0.00, 0.11]
QSKINI 0.05 [0.03, 0.08]
QSKINII 0.04 [0.00, 0.08]
AGDS 0.02 [-0.01, 0.05]
-0.4 -0.2 0 0.2 0.4
Standardized effect of 2:38298139*T

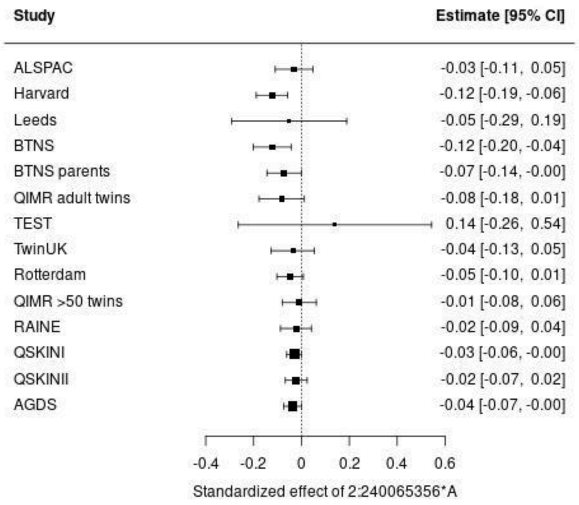
Study
Estimate [95% CI]
ALSPAC -0.03 [-0.11, 0.05]
Harvard -0.12 [-0.19, -0.06]
Leeds -0.05 [-0.29, 0.19]
BTNS -0.12 [-0.20, -0.04]
BTNS parents -0.07 [-0.14, -0.00]
QIMR adult twins -0.08 [-0.18, 0.01]
TEST 0.14 [-0.26, 0.54]
TwinUK -0.04 [-0.13, 0.05]
Rotterdam -0.05 [-0.10, 0.01]
QIMR >50 twins -0.01 [-0.08, 0.06]
RAINE -0.02 [-0.09, 0.04]
QSKINI -0.03 [-0.06, -0.00]
QSKINII -0.02 [-0.07, 0.02]
AGDS -0.04 [-0.07, -0.00]
-0.4 -0.2 0 0.2 0.4 0.6
Standardized effect of 2:240065356*A

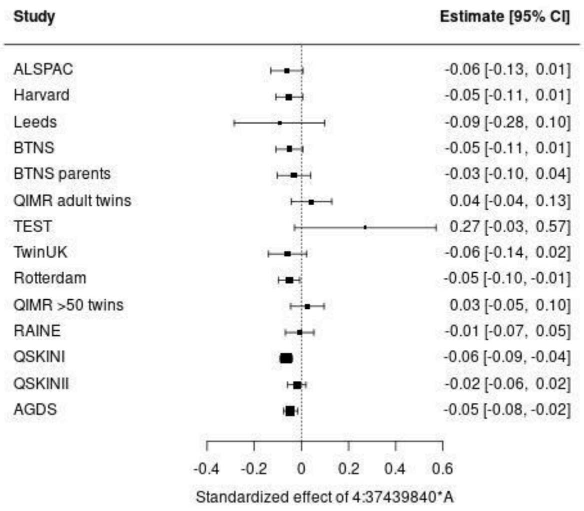
Study
Estimate [95% CI]
ALSPAC -0.06 [-0.13, 0.01]
Harvard -0.05 [-0.11, 0.01]
Leeds -0.09 [-0.28, 0.10]
BTNS -0.05 [-0.11, 0.01]
BTNS parents -0.03 [-0.10, 0.04]
QIMR adult twins 0.04 [-0.04, 0.13]
TEST 0.27 [-0.03, 0.57]
TwinUK -0.06 [-0.14, 0.02]
Rotterdam -0.05 [-0.10, -0.01]
QIMR >50 twins 0.03 [-0.05, 0.10]
RAINE -0.01 [-0.07, 0.05]
QSKINI -0.06 [-0.09, -0.04]
QSKINII -0.02 [-0.06, 0.02]
AGDS -0.05 [-0.08, -0.02]
-0.4 -0.2 0 0.2 0.4 0.6
Standardized effect of 4:37439840*A

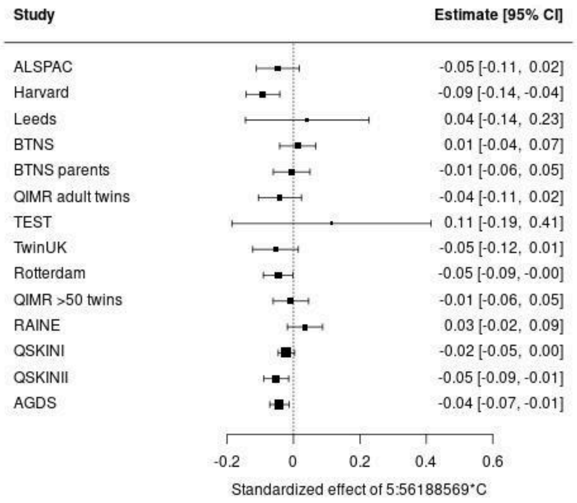
Study
Estimate [95% CI]
ALSPAC -0.05 [-0.11, 0.02]
Harvard -0.09 [-0.14, -0.04]
Leeds 0.04 [-0.14, 0.23]
BTNS 0.01 [-0.04, 0.07]
BTNS parents -0.01 [-0.06, 0.05]
QIMR adult twins -0.04 [-0.11, 0.02]
TEST 0.11 [-0.19, 0.41]
TwinUK -0.05 [-0.12, 0.01]
Rotterdam -0.05 [-0.09, -0.00]
QIMR >50 twins -0.01 [-0.06, 0.05]
RAINE 0.03 [-0.02, 0.09]
QSKINI -0.02 [-0.05, 0.00]
QSKINII -0.05 [-0.09, -0.01]
AGDS -0.04 [-0.07, -0.01]
-0.2 0 0.2 0.4 0.6
Standardized effect of 5:56188569*C

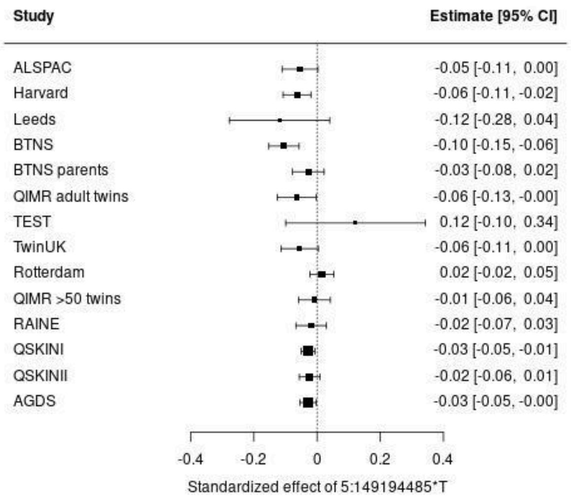
Study
Estimate [95% CI]
ALSPAC -0.05 [-0.11, 0.00]
Harvard -0.06 [-0.11, -0.02]
Leeds -0.12 [-0.28, 0.04]
BTNS -0.10 [-0.15, -0.06]
BTNS parents -0.03 [-0.08, 0.02]
QIMR adult twins -0.06 [-0.13, -0.00]
TEST 0.12 [-0.10, 0.34]
TwinUK -0.06 [-0.11, 0.00]
Rotterdam 0.02 [-0.02, 0.05]
QIMR >50 twins -0.01 [-0.06, 0.04]
RAINE -0.02 [-0.07, 0.03]
QSKINI -0.03 [-0.05, -0.01]
QSKINII -0.02 [-0.06, 0.01]
AGDS -0.03 [-0.05, -0.00]
-0.4 -0.2 0 0.2 0.4
Standardized effect of 5:149194485*T

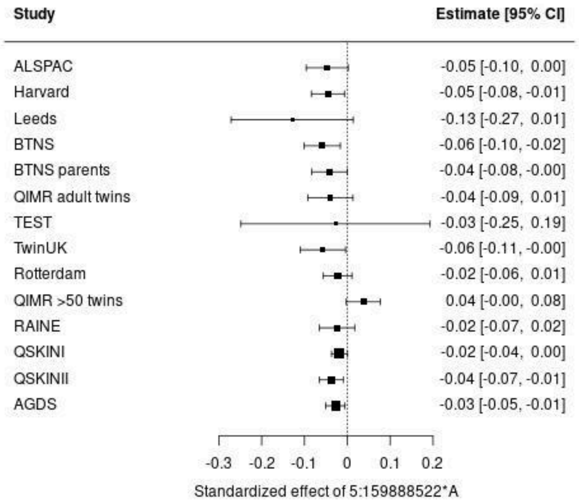
Study
Estimate [95% CI]
ALSPAC -0.05 [-0.10, 0.00]
Harvard -0.05 [-0.08, -0.01]
Leeds -0.13 [-0.27, 0.01]
BTNS -0.06 [-0.10, -0.02]
BTNS parents -0.04 [-0.08, -0.00]
QIMR adult twins -0.04 [-0.09, 0.01]
TEST -0.03 [-0.25, 0.19]
TwinUK -0.06 [-0.11, -0.00]
Rotterdam -0.02 [-0.06, 0.01]
QIMR >50 twins 0.04 [-0.00, 0.08]
RAINE -0.02 [-0.07, 0.02]
QSKINI -0.02 [-0.04, 0.00]
QSKINII -0.04 [-0.07, -0.01]
AGDS -0.03 [-0.05, -0.01]
-0.3 -0.2 -0.1 0 0.1 0.2
Standardized effect of 5:159888522*A

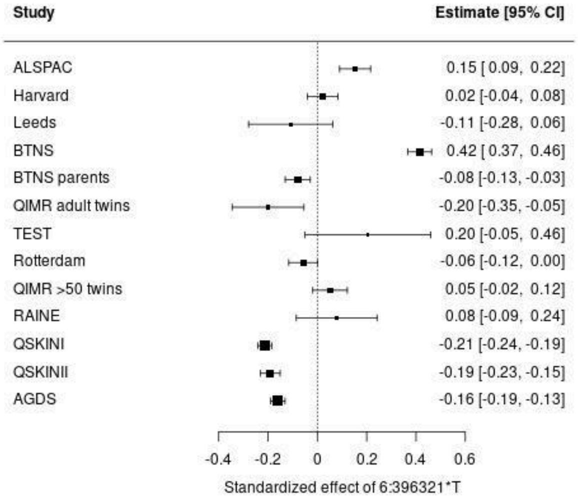
Study
Estimate [95% CI]
ALSPAC 0.15 [0.09, 0.22]
Harvard 0.02 [-0.04, 0.08]
Leeds -0.11 [-0.28, 0.06]
BTNS 0.42 [0.37, 0.46]
BTNS parents -0.08 [-0.13, -0.03]
QIMR adult twins -0.20 [-0.35, -0.05]
TEST 0.20 [-0.05, 0.46]
Rotterdam -0.06 [-0.12, 0.00]
QIMR >50 twins 0.05 [-0.02, 0.12]
RAINE 0.08 [-0.09, 0.24]
QSKINI -0.21 [-0.24, -0.19]
QSKINII -0.19 [-0.23, -0.15]
AGDS -0.16 [-0.19, -0.13]
-0.4 -0.2 0 0.2 0.4 0.6
Standardized effect of 6:396321*T

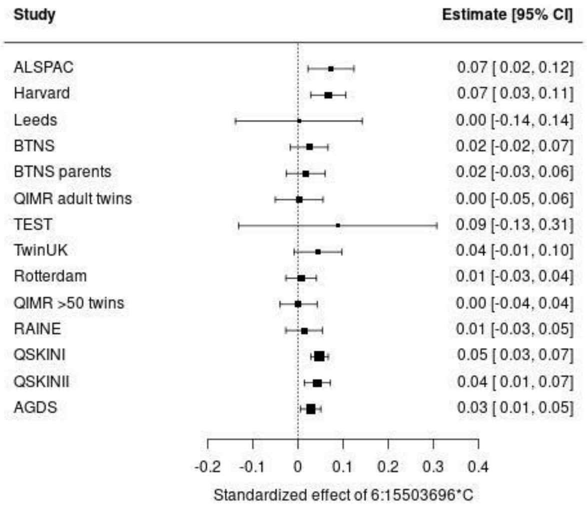
Study
Estimate [95% CI]
ALSPAC 0.07 [0.02, 0.12]
Harvard 0.07 [0.03, 0.11]
Leeds 0.00 [-0.14, 0.14]
BTNS 0.02 [-0.02, 0.07]
BTNS parents 0.02 [-0.03, 0.06]
QIMR adult twins 0.00 [-0.05, 0.06]
TEST 0.09 [-0.13, 0.31]
TwinUK 0.04 [-0.01, 0.10]
Rotterdam 0.01 [-0.03, 0.04]
QIMR >50 twins 0.00 [-0.04, 0.04]
RAINE 0.01 [-0.03, 0.05]
QSKINI 0.05 [0.03, 0.07]
QSKINII 0.04 [0.01, 0.07]
AGDS 0.03 [0.01, 0.05]
-0.2 -0.1 0 0.1 0.2 0.3 0.4
Standardized effect of 6:15503696*C

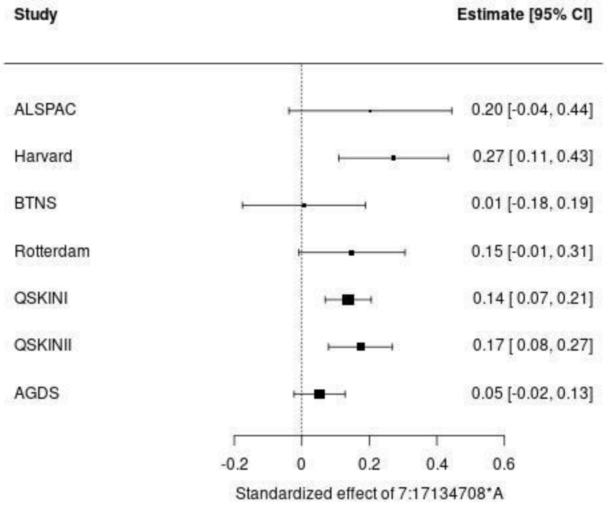
Study
Estimate [95% CI]
ALSPAC 0.20 [-0.04, 0.44]
Harvard 0.27 [0.11, 0.43]
BTNS 0.01 [-0.18, 0.19]
Rotterdam 0.15 [-0.01, 0.31]
QSKINI 0.14 [0.07, 0.21]
QSKINII 0.17 [0.08, 0.27]
AGDS 0.05 [-0.02, 0.13]
-0.2 0 0.2 0.4 0.6
Standardized effect of 7:17134708*A

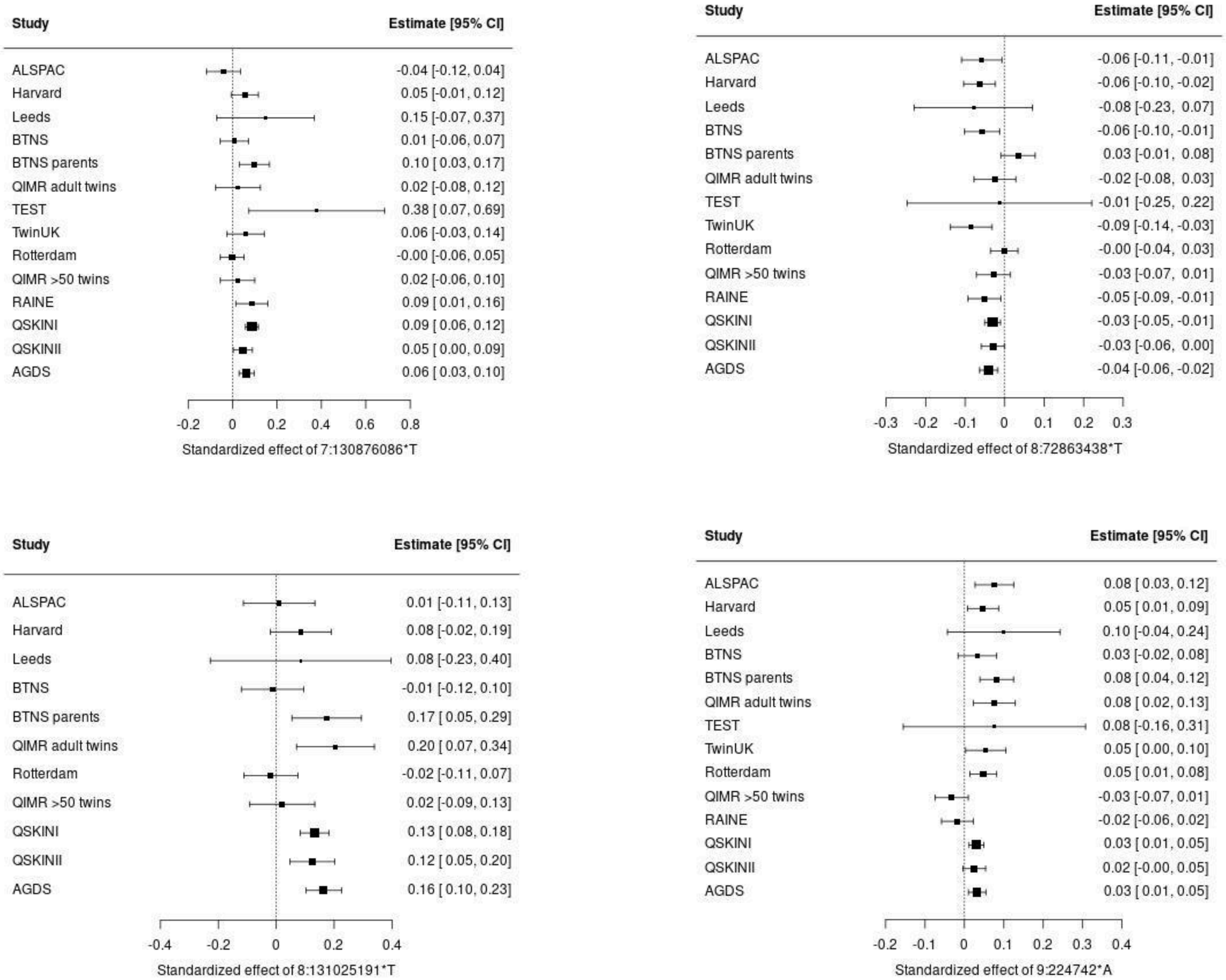
Study
Estimate [95% CI]
ALSPAC -0.04 [-0.12, 0.04]
Harvard 0.05 [-0.01, 0.12]
Leeds 0.15 [-0.07, 0.37]
BTNS 0.01 [-0.06, 0.07]
BTNS parents 0.10 [ 0.03, 0.17]
QIMR adult twins 0.02 [-0.08, 0.12]
TEST 0.38 [ 0.07, 0.69]
TwinUK 0.06 [-0.03, 0.14]
Rotterdam -0.00 [-0.06, 0.05]
QIMR >50 twins 0.02 [-0.06, 0.10]
RAINE 0.09 [ 0.01, 0.16]
QSKINI 0.09 [ 0.06, 0.12]
QSKINII 0.05 [ 0.00, 0.09]
AGDS 0.06 [ 0.03, 0.10]
-0.2 0 0.2 0.4 0.6 0.8
Standardized effect of 7:130876086*T
Study
Estimate [95% CI]
ALSPAC -0.06 [-0.11, -0.01]
Harvard -0.06 [-0.10, -0.02]
Leeds -0.08 [-0.23, 0.07]
BTNS -0.06 [-0.10, -0.01]
BTNS parents 0.03 [-0.01, 0.08]
QIMR adult twins -0.02 [-0.08, 0.03]
TEST -0.01 [-0.25, 0.22]
TwinUK -0.09 [-0.14, -0.03]
Rotterdam -0.00 [-0.04, 0.03]
QIMR >50 twins -0.03 [-0.07, 0.01]
RAINE -0.05 [-0.09, -0.01]
QSKINI -0.03 [-0.05, -0.01]
QSKINII -0.03 [-0.06, 0.00]
AGDS -0.04 [-0.06, -0.02]
-0.3 -0.2 -0.1 0 0.1 0.2 0.3
Standardized effect of 8:72863438*T
Study
Estimate [95% CI]
ALSPAC 0.01 [-0.11, 0.13]
Harvard 0.08 [-0.02, 0.19]
Leeds 0.08 [-0.23, 0.40]
BTNS -0.01 [-0.12, 0.10]
BTNS parents 0.17 [ 0.05, 0.29]
QIMR adult twins 0.20 [ 0.07, 0.34]
Rotterdam -0.02 [-0.11, 0.07]
QIMR >50 twins 0.02 [-0.09, 0.13]
QSKINI 0.13 [ 0.08, 0.18]
QSKINII 0.12 [ 0.05, 0.20]
AGDS 0.16 [ 0.10, 0.23]
-0.4 -0.2 0 0.2 0.4
Standardized effect of 8:131025191*T
Study
Estimate [95% CI]
ALSPAC 0.08 [ 0.03, 0.12]
Harvard 0.05 [ 0.01, 0.09]
Leeds 0.10 [-0.04, 0.24]
BTNS 0.03 [-0.02, 0.08]
BTNS parents 0.08 [ 0.04, 0.12]
QIMR adult twins 0.08 [ 0.02, 0.13]
TEST 0.08 [-0.16, 0.31]
TwinUK 0.05 [ 0.00, 0.10]
Rotterdam 0.05 [ 0.01, 0.08]
QIMR >50 twins -0.03 [-0.07, 0.01]
RAINE -0.02 [-0.06, 0.02]
QSKINI 0.03 [ 0.01, 0.05]
QSKINII 0.02 [-0.00, 0.05]
AGDS 0.03 [ 0.01, 0.05]
-0.2 -0.1 0 0.1 0.2 0.3 0.4
Standardized effect of 9:224742*A

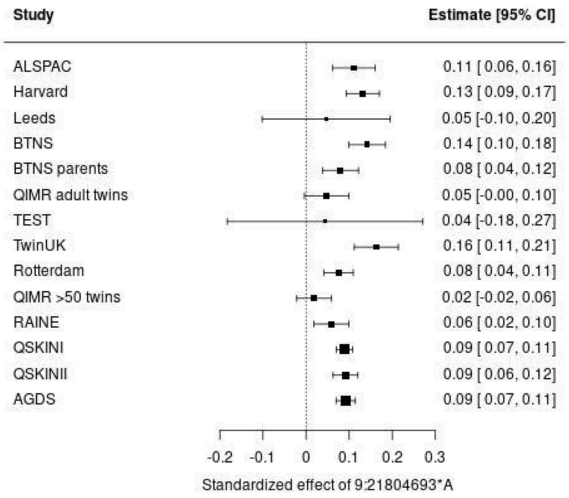
Study
Estimate [95% CI]
ALSPAC 0.11 [ 0.06, 0.16]
Harvard 0.13 [ 0.09, 0.17]
Leeds 0.05 [-0.10, 0.20]
BTNS 0.14 [ 0.10, 0.18]
BTNS parents 0.08 [ 0.04, 0.12]
QIMR adult twins 0.05 [-0.00, 0.10]
TEST 0.04 [-0.18, 0.27]
TwinUK 0.16 [ 0.11, 0.21]
Rotterdam 0.08 [ 0.04, 0.11]
QIMR >50 twins 0.02 [-0.02, 0.06]
RAINE 0.06 [ 0.02, 0.10]
QSKINI 0.09 [ 0.07, 0.11]
QSKINII 0.09 [ 0.06, 0.12]
AGDS 0.09 [ 0.07, 0.11]
-0.2 -0.1 0 0.1 0.2 0.3
Standardized effect of 9:21804693*A

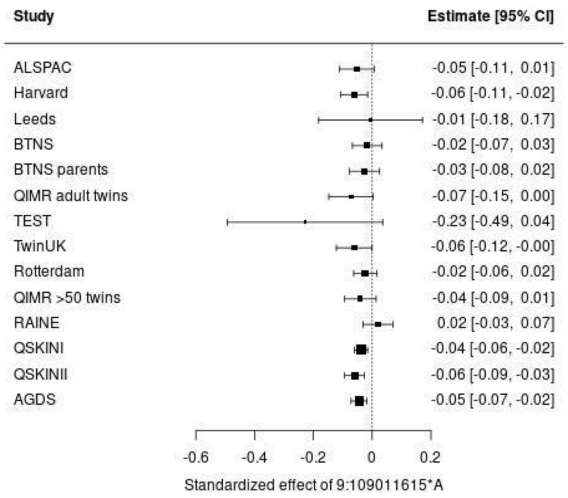
Study
Estimate [95% CI]
ALSPAC -0.05 [-0.11, 0.01]
Harvard -0.06 [-0.11, -0.02]
Leeds -0.01 [-0.18, 0.17]
BTNS -0.02 [-0.07, 0.03]
BTNS parents -0.03 [-0.08, 0.02]
QIMR adult twins -0.07 [-0.15, 0.00]
TEST -0.23 [-0.49, 0.04]
TwinUK -0.06 [-0.12, -0.00]
Rotterdam -0.02 [-0.06, 0.02]
QIMR >50 twins -0.04 [-0.09, 0.01]
RAINE 0.02 [-0.03, 0.07]
QSKINI -0.04 [-0.06, -0.02]
QSKINII -0.06 [-0.09, -0.03]
AGDS -0.05 [-0.07, -0.02]
-0.6 -0.4 -0.2 0 0.2
Standardized effect of 9:109011615*A

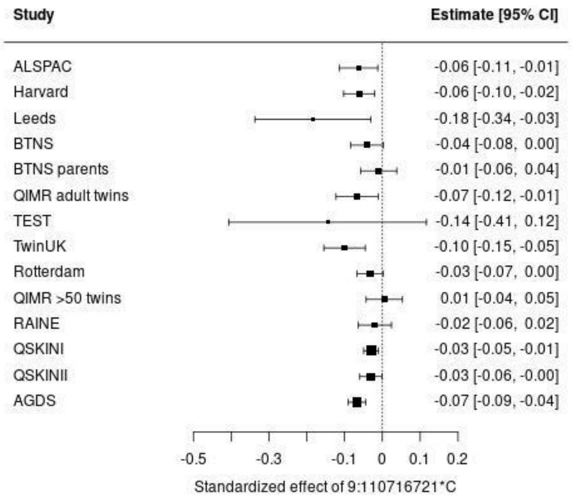
Study
Estimate [95% CI]
ALSPAC -0.06 [-0.11, -0.01]
Harvard -0.06 [-0.10, -0.02]
Leeds -0.18 [-0.34, -0.03]
BTNS -0.04 [-0.08, 0.00]
BTNS parents -0.01 [-0.06, 0.04]
QIMR adult twins -0.07 [-0.12, -0.01]
TEST -0.14 [-0.41, 0.12]
TwinUK -0.10 [-0.15, -0.05]
Rotterdam -0.03 [-0.07, 0.00]
QIMR >50 twins 0.01 [-0.04, 0.05]
RAINE -0.02 [-0.06, 0.02]
QSKINI -0.03 [-0.05, -0.01]
QSKINII -0.03 [-0.06, -0.00]
AGDS -0.07 [-0.09, -0.04]
-0.5 -0.3 -0.1 0 0.1 0.2
Standardized effect of 9:110716721*C

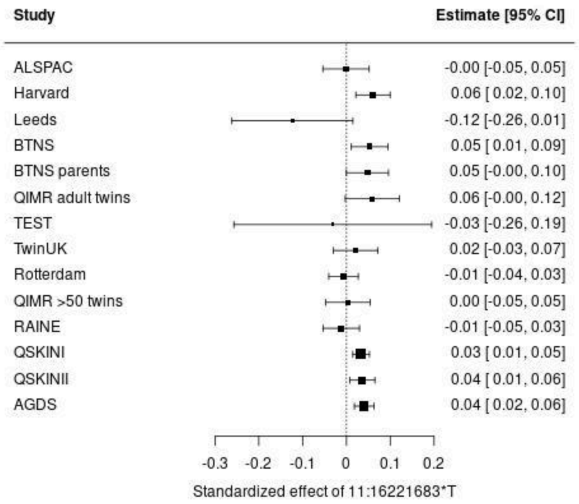
Study
Estimate [95% CI]
ALSPAC -0.00 [-0.05, 0.05]
Harvard 0.06 [ 0.02, 0.10]
Leeds -0.12 [-0.26, 0.01]
BTNS 0.05 [ 0.01, 0.09]
BTNS parents 0.05 [-0.00, 0.10]
QIMR adult twins 0.06 [-0.00, 0.12]
TEST -0.03 [-0.26, 0.19]
TwinUK 0.02 [-0.03, 0.07]
Rotterdam -0.01 [-0.04, 0.03]
QIMR >50 twins 0.00 [-0.05, 0.05]
RAINE -0.01 [-0.05, 0.03]
QSKINI 0.03 [ 0.01, 0.05]
QSKINII 0.04 [ 0.01, 0.06]
AGDS 0.04 [ 0.02, 0.06]
-0.3 -0.2 -0.1 0 0.1 0.2
Standardized effect of 11:16221683*T

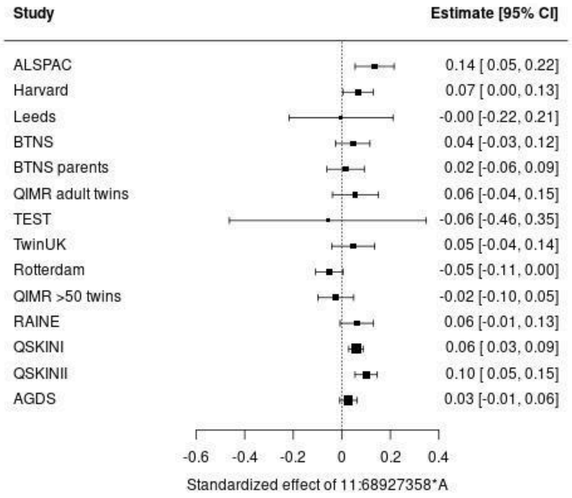
Study
Estimate [95% CI]
ALSPAC 0.14 [ 0.05, 0.22]
Harvard 0.07 [ 0.00, 0.13]
Leeds -0.00 [-0.22, 0.21]
BTNS 0.04 [-0.03, 0.12]
BTNS parents 0.02 [-0.06, 0.09]
QIMR adult twins 0.06 [-0.04, 0.15]
TEST -0.06 [-0.46, 0.35]
TwinUK 0.05 [-0.04, 0.14]
Rotterdam -0.05 [-0.11, 0.00]
QIMR >50 twins -0.02 [-0.10, 0.05]
RAINE 0.06 [-0.01, 0.13]
QSKINI 0.06 [ 0.03, 0.09]
QSKINII 0.10 [ 0.05, 0.15]
AGDS 0.03 [-0.01, 0.06]
-0.6 -0.4 -0.2 0 0.2 0.4
Standardized effect of 11:68927358*A

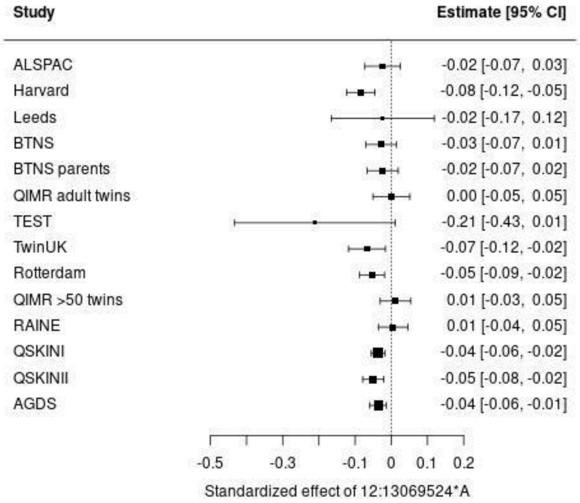
Study
Estimate [95% CI]
ALSPAC -0.02 [-0.07, 0.03]
Harvard -0.08 [-0.12, -0.05]
Leeds -0.02 [-0.17, 0.12]
BTNS -0.03 [-0.07, 0.01]
BTNS parents -0.02 [-0.07, 0.02]
QIMR adult twins 0.00 [-0.05, 0.05]
TEST -0.21 [-0.43, 0.01]
TwinUK -0.07 [-0.12, -0.02]
Rotterdam -0.05 [-0.09, -0.02]
QIMR >50 twins 0.01 [-0.03, 0.05]
RAINE 0.01 [-0.04, 0.05]
QSKINI -0.04 [-0.06, -0.02]
QSKINII -0.05 [-0.08, -0.02]
AGDS -0.04 [-0.06, -0.01]
-0.5 -0.3 -0.1 0 0.1 0.2
Standardized effect of 12:13069524*A

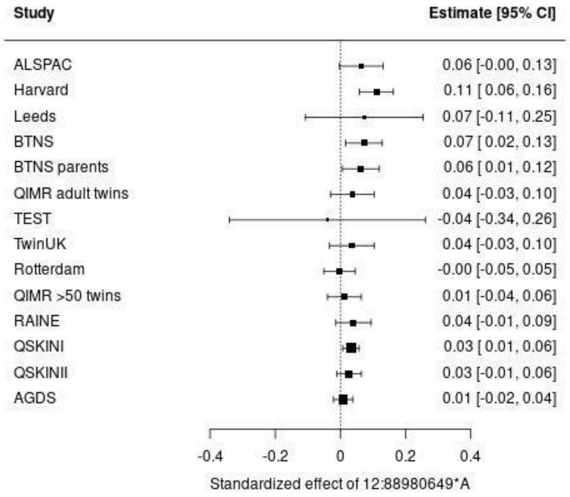
Study
Estimate [95% CI]
ALSPAC 0.06 [-0.00, 0.13]
Harvard 0.11 [ 0.06, 0.16]
Leeds 0.07 [-0.11, 0.25]
BTNS 0.07 [ 0.02, 0.13]
BTNS parents 0.06 [ 0.01, 0.12]
QIMR adult twins 0.04 [-0.03, 0.10]
TEST -0.04 [-0.34, 0.26]
TwinUK 0.04 [-0.03, 0.10]
Rotterdam -0.00 [-0.05, 0.05]
QIMR >50 twins 0.01 [-0.04, 0.06]
RAINE 0.04 [-0.01, 0.09]
QSKINI 0.03 [ 0.01, 0.06]
QSKINII 0.03 [-0.01, 0.06]
AGDS 0.01 [-0.02, 0.04]
-0.4 -0.2 0 0.2 0.4
Standardized effect of 12:88980649*A

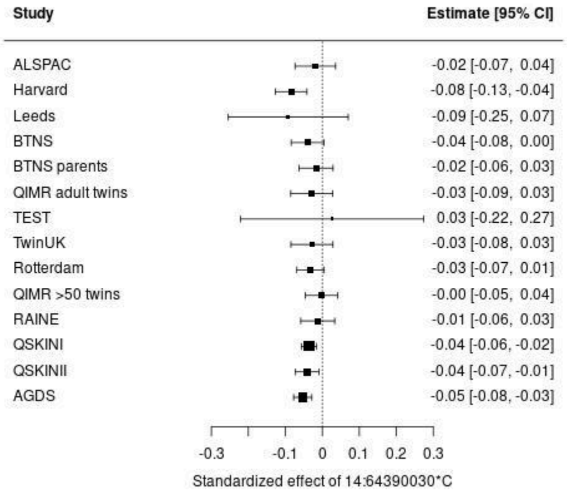
Study
Estimate [95% CI]
ALSPAC -0.02 [-0.07, 0.04]
Harvard -0.08 [-0.13, -0.04]
Leeds -0.09 [-0.25, 0.07]
BTNS -0.04 [-0.08, 0.00]
BTNS parents -0.02 [-0.06, 0.03]
QIMR adult twins -0.03 [-0.09, 0.03]
TEST 0.03 [-0.22, 0.27]
TwinUK -0.03 [-0.08, 0.03]
Rotterdam -0.03 [-0.07, 0.01]
QIMR >50 twins -0.00 [-0.05, 0.04]
RAINE -0.01 [-0.06, 0.03]
QSKINI -0.04 [-0.06, -0.02]
QSKINII -0.04 [-0.07, -0.01]
AGDS -0.05 [-0.08, -0.03]
-0.3 -0.1 0 0.1 0.2 0.3
Standardized effect of 14:64390030*C

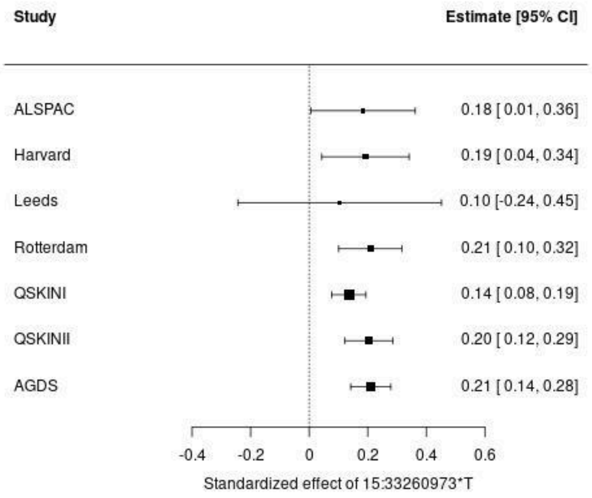
Study
Estimate [95% CI]
ALSPAC 0.18 [ 0.01, 0.36]
Harvard 0.19 [ 0.04, 0.34]
Leeds 0.10 [-0.24, 0.45]
Rotterdam 0.21 [ 0.10, 0.32]
QSKINI 0.14 [ 0.08, 0.19]
QSKINII 0.20 [ 0.12, 0.29]
AGDS 0.21 [ 0.14, 0.28]
-0.4 -0.2 0 0.2 0.4 0.6
Standardized effect of 15:33260973*T

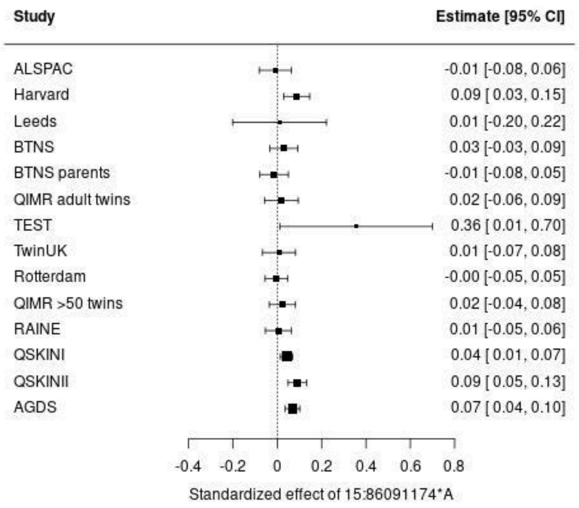
Study
Estimate [95% CI]
ALSPAC -0.01 [-0.08, 0.06]
Harvard 0.09 [ 0.03, 0.15]
Leeds 0.01 [-0.20, 0.22]
BTNS 0.03 [-0.03, 0.09]
BTNS parents -0.01 [-0.08, 0.05]
QIMR adult twins 0.02 [-0.06, 0.09]
TEST 0.36 [ 0.01, 0.70]
TwinUK 0.01 [-0.07, 0.08]
Rotterdam -0.00 [-0.05, 0.05]
QIMR >50 twins 0.02 [-0.04, 0.08]
RAINE 0.01 [-0.05, 0.06]
QSKINI 0.04 [ 0.01, 0.07]
QSKINII 0.09 [ 0.05, 0.13]
AGDS 0.07 [ 0.04, 0.10]
-0.4 -0.2 0 0.2 0.4 0.6 0.8
Standardized effect of 15:86091174*A

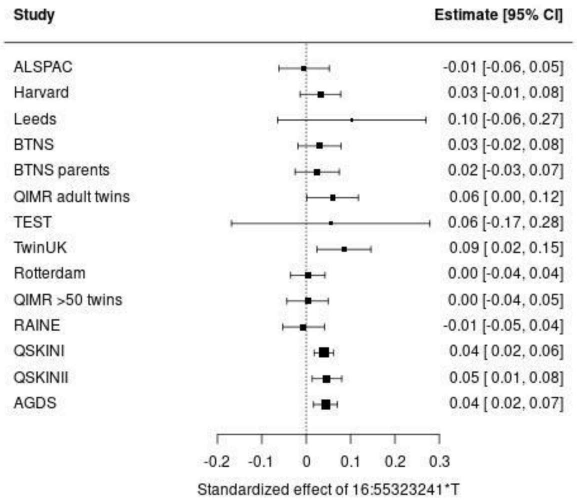
Study
Estimate [95% CI]
ALSPAC -0.01 [-0.06, 0.05]
Harvard 0.03 [-0.01, 0.08]
Leeds 0.10 [-0.06, 0.27]
BTNS 0.03 [-0.02, 0.08]
BTNS parents 0.02 [-0.03, 0.07]
QIMR adult twins 0.06 [ 0.00, 0.12]
TEST 0.06 [-0.17, 0.28]
TwinUK 0.09 [ 0.02, 0.15]
Rotterdam 0.00 [-0.04, 0.04]
QIMR >50 twins 0.00 [-0.04, 0.05]
RAINE -0.01 [-0.05, 0.04]
QSKINI 0.04 [ 0.02, 0.06]
QSKINII 0.05 [ 0.01, 0.08]
AGDS 0.04 [ 0.02, 0.07]
-0.2 -0.1 0 0.1 0.2 0.3
Standardized effect of 16:55323241*T

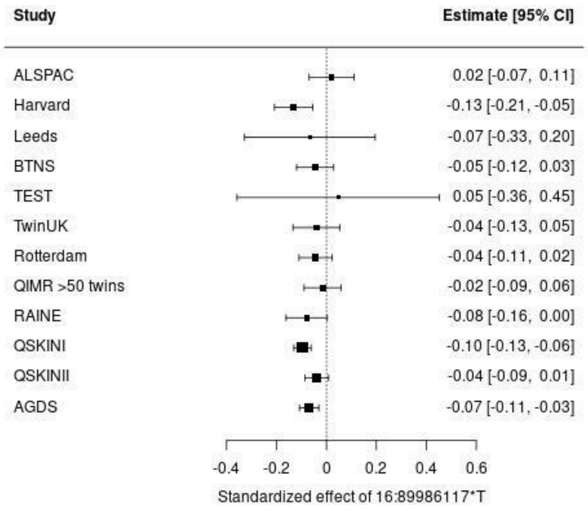
Study
Estimate [95% CI]
ALSPAC 0.02 [-0.07, 0.11]
Harvard -0.13 [-0.21, -0.05]
Leeds -0.07 [-0.33, 0.20]
BTNS -0.05 [-0.12, 0.03]
TEST 0.05 [-0.36, 0.45]
TwinUK -0.04 [-0.13, 0.05]
Rotterdam -0.04 [-0.11, 0.02]
QIMR >50 twins -0.02 [-0.09, 0.06]
RAINE -0.08 [-0.16, 0.00]
QSKINI -0.10 [-0.13, -0.06]
QSKINII -0.04 [-0.09, 0.01]
AGDS -0.07 [-0.11, -0.03]
-0.4 -0.2 0 0.2 0.4 0.6
Standardized effect of 16:89986117*T

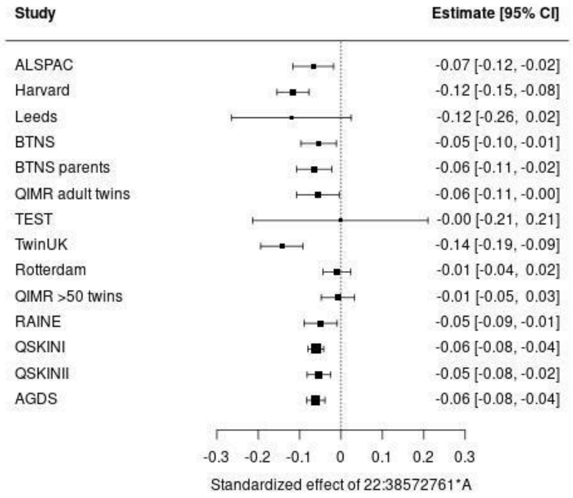


Supplementary Figure 18: Forest plot for 29 significant genomic loci associated with nevus count identified through GWAS meta-analysis conducted using all 14 GWAS

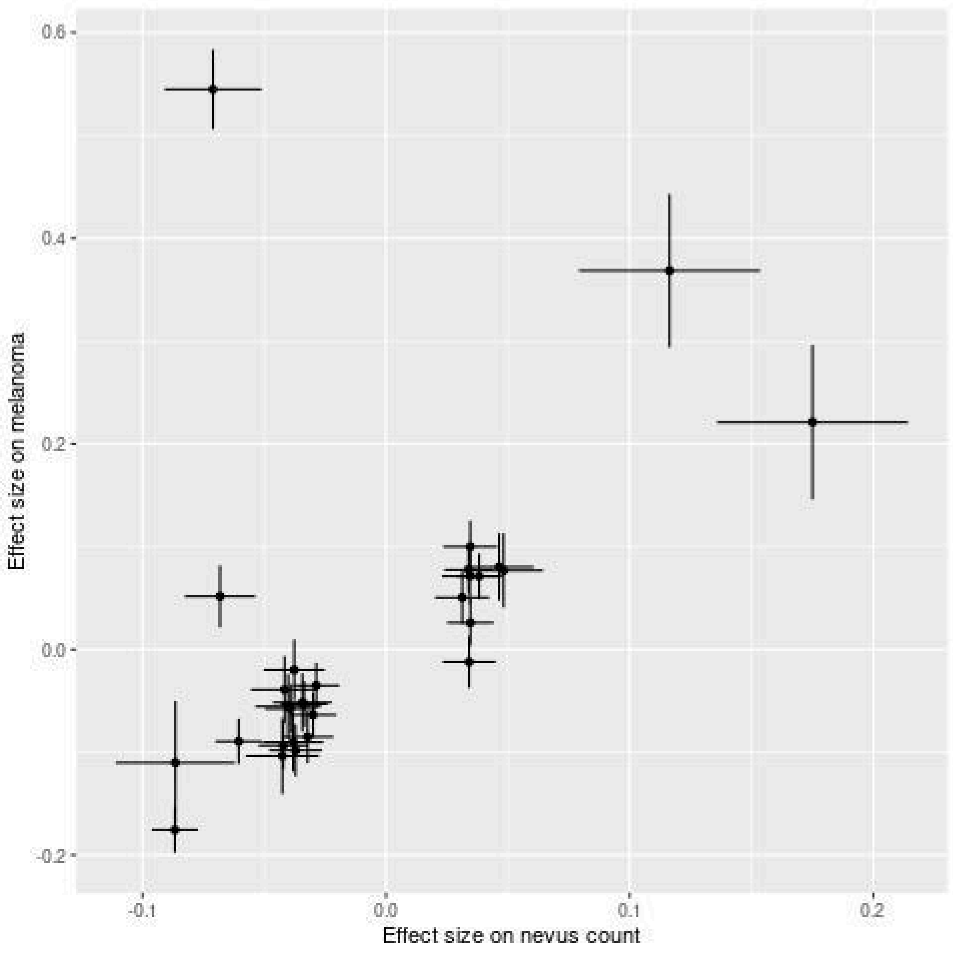


Supplementary Figure 19: Plot of effect size of effect alleles of significant-top SNPs on 29 nevus loci for nevus count and for melanoma from published melanoma GWAS[14]

**Supplementary Notes S1 Description of study cohorts used for nevus GWAS meta-analysis**

***S1.1* Study cohorts from previously published nevus GWAS meta-analysis**[1]

Our meta-analysis was based on 14 GWAS studies of nevus count, conducted on samples of healthy individuals without melanoma of European ancestry. Eleven of those 14 GWASs have been previously published and comprehensive details on participant recruitment, phenotype measurement and genotyping of these study cohorts have been reported elsewhere[1] and briefly outlined in Table S1.1. Nevus count assessments varied significantly across studies due to differences in the anatomical site where measurement has been obtained, the size threshold of the nevi, the counting method; self-counted or counted by observer, and the scale; categorical or numerical count.

Nevus meta-analysis conducted by[1] has included samples with related individuals (e.g. BTNS, Parents of BTNS and TwinsUK) or unrelated individuals (Table S1.1). Therefore, for samples with related individuals, MERLIN or MENDEL has been used to test the association between GWAS SNPs and nevus count while accounting for relatedness, whereas unrelated individuals have been analysed in PLINK[1]. In each analysis, sex, age, $age^2$, age x sex, $age^2$ x sex and up to 5 PCs were included as covariates[1]

Table S1.1: Details of published GWAS studies from[1]

| Study | SNP chip | GWAS sample size | Description of nevus count |
|---|---|---|---|
| Avon Longitudinal Study of Parents and Children (ALSPAC) in UK (Bristol)[2] | 550k | 3,309 | Self-counted large and small nevi on the arms and legs |
| Harvard<br>1. Nurses' Health Study (NHS)<br>2. Nurses' Health Study II (NHS2)<br>3. Health Professionals Follow-up Study (HPFS)<br>in US (Boston)[3] | Affy+Illumina various | 32,975 | Self-counted nevi (> 3 mm)<br>on the arms (Categories: 1 = none, 2 = 1–2, 3 = 3–5, 4 = 6–9, 5 = 10–14, 6 = 15–20, and 7 = 21+) |
| Leeds in Yorkshire[4] | OmniExpressExome | 397 | Nurse-counted total body nevus count (>2 mm) |
| QIMR Brisbane Twin Nevus Study (BTNS) in Brisbane[5] | 610k, CoreExome | 3,261 (1,309 families) | Nurse-counted total body nevus count (>0 mm) |
| Parents of BTNS Twins in Southeastern Queensland[6] | 610k+CoreExome | 2,248 (families 1,299) | Self-rating total body 4-point scale: none, a few, moderate, many (item was accompanied by a<br>cartoon showing 0, 6, 14 and 28 spots ) |
| QIMR Adult Twins in Australia[7] | 317k+370k+610k+ CE | 1,848 (families 1,113) | Self-rating 4-point scale total body nevus count: (1) None, (2) <10, (3)10–50, (4) >50 |
| QIMR >50 Twins in Australia[8] | 370k+610k+CE | 893 (families 596) | Self-counted nevi (>4 mm) on right arm |
| The Raine Study in Western Australia[9] | 660k | 808 | Research assistant-counted nevi (<2 mm diameter, 2–5 mm, >5 mm) on the right arm |

| | | | |
|---|---|---|---|
| Rotterdam Study in Rotterdam[10] | 550k, 610k | 3319 | Dermatology resident-counted total body nevus count (>2 mm diameter) 4-point scale: <25, 25–50, 50–100, >100 |
| Twins Eyes Study in Tasmania (TEST)[11] | 610k+CE | 136 (71 families) | Nurse-counted total body nevus count (>0 mm) |
| St. Thomas' UK Adult Twin Registry (TwinsUK) Study in London[12] | 317k+610k+1M+ 1.2M | 3312 (1839 families) | Nurse-counted total body nevus count (>2 mm) |

### *S1.2 New nevus study cohorts*

**QSkin Sun and Health Study Phase I (QSkin I)**

The QSkin Sun and Health Study is a population-based cohort of over 43,000 participants recruited from Queensland, Australia in 2011[13]. This study focused on exploring the impact of environmental and host/genetic factors in the development of cutaneous melanoma and other skin cancers. Comprehensive details regarding participant recruitment, genotyping, imputation and QC procedures can be found elsewhere[13,14]. In brief, 19,119 participants were genotyped using the Global Screening Array [Illumina, San Diego, USA]. Before imputation, exclusion criteria for individuals involved non-European ancestry (beyond 6 standard deviations (SD) from the mean of principal components (PC) 1 and PC2 of 1000 Genomes European samples), relatedness (identify-by-descent pihat > 0.1875), and high genotype missingness (> 3%). SNPs were filtered out due to call rate below 97%, HWE failure (P-value $< 1 \times 10^{-6}$), low GenTrain score (< 0.6), or low MAF (<1%). Cleaned genotype data were imputed to the Haplotype Reference Consortium (HRC) version 1 panel via the University of Michigan imputation server[15,16]. Through questionnaires, participants reported total body nevus count on a 4-point scale with categories: none, few, moderate, and many, illustrated by a pictogram depicting spots on a human figure. In addition, nevus count (nevi > 2 mm) on the left upper arm was also collected from the participants. For categorical nevus phenotype, nevus data and quality-controlled genotypes were available for 15,348 individuals while for absolute nevus count data, the sample size was 13,007. In both samples, related individuals were retained, because, in genome-wide association analysis we have used SAIGE (Scalable and Accurate Implementation of GEneralized mixed model)[17] which effectively incorporates relatedness.

**QSkin Sun and Health Study Phase II (QSkin II)**

This is the second phase of the QSkin Sun and Health Study. The same protocols as implemented in QSkin I, were applied for participant recruitment, phenotype measurement, genotyping, and QC procedures. Initially, genotype data was available for 8,296 participants and for the genetic association study, 4-point scaled nevus count data and quality-controlled genotypes were available for 6,608 individuals including related individuals.

**Australian Genetics of Depression Study (AGDS)**

AGDS cohort was established to explore genetic and environmental factors influencing depression and response to common antidepressants. 20,689 participants were recruited and comprised predominantly female (75%), with an average age of 43[18]. Genotyping was conducted using Illumina Global Screening Array V.2.0 (GSA) and cleaned by removing ambiguous, high missingness (> 5%), deviation from HWE (P-value $< 1 \times 10^{-6}$), low MAF (< 1%), and GenTrain score < 0.6 variants, and participants with non-European ancestry before imputation through the Michigan imputation server web service using the HRCr1.1 reference panel[15,16]. Further details on participant recruitment and genotyping have been published elsewhere[18]. The total body count was recorded by participants based on a 4-point scale: (1) None, (2) <10, (3) 10–50, (4) >50 and genotypes filtered for quality control with nevus count data were available for 11,775 individuals. Related individuals were retained here since we used SAIGE in genome-wide association analysis.

**GWAS for QSkin I, QSkin II and AGDS**

A subset of individuals in QSKIN I have self-reported 4-point whole-body nevus count (none, some, few, many) and also reported nevus count on their upper arms. We reanalysed QSkin I following the inverse-normal rank transformation applied to 4-point scale nevus count ($n$ = 15,348) considered as a quantitative trait, and nevus count ($n$ = 13,007) on the upper arm using the SAIGE method [17]. Then we analysed the untransformed 4-point scale using the POLMM [19] Among SAIGE and POLMM, the most powerful approach was selected based on the identified number of significant loci and estimated SNP-based heritability using LDSC [20] as explained below. By applying each SAIGE method and POLMM method for 4-point scale nevus count, six significant genomic loci (P < 5e-8) were identified, and the majority of them were overlapped in both methods (Supplementary Figure 1-2). However, SNP-based heritability was slightly higher for SAIGE method ($h^2$ = 0.15, SE = 0.038) compared to POLMM method ($h^2$ = 0.13, SE = 0.036). Considering both facts, we selected SAIGE method for conducting GWAS on a 4-point scale nevus count of QSkin II and AGDS. In addition, upper arm nevus count GWAS of QSkin I found only two significant genomic loci (P < 5e-8, Supplementary Figure 3) and GWAS SNP-based heritability was 0.16 (SE = 0.044). The 4-point scale nevus count GWAS of QSkin I was used for further analysis since it found more significant loci compared to upper arm nevus count GWAS, indicating a greater power. In each GWAS, the inverse-normal rank transformation was applied to the nevus count when applying SAIGE method, and sex, the orthogonal transformation of age, age×sex, $age^2$×sex and the first 10 PCs used to account for any residual population stratification were considered covariates. In addition to these covariates, in QSkin I, a batch variable is added to account for the batch effect since the genotyping for QSkin I was conducted in two batches. For downstream analysis, variants in each GWAS were further filtered for imputation quality score > 0.5 and MAF > 0.005.

**Acknowledgments for contributing studies**
**Nevus GWAS from the published meta-analysis**[1]
Full details of financial support and acknowledgements are given in [1] and a summary is included here. Along with the specific acknowledgments listed below, we also extend our gratitude to G. Clement, Bernet Keto, Pirro Hysi, and Emad Qweitin for their support.

**ALSPAC**: This study was supported by the UK Medical Research Council, Wellcome Trust (Grant ref: 102215/2/13/2), the University of Bristol, Sample Logistics and Genotyping Facilities at the Wellcome Trust Sanger Institute and LabCorp (Laboratory Corporation of America). 23andMe, ARC Future Fellowship (FT130101709), and a Medical Research Council program grant (MC_UU_12013/4).

**Harvard**: We are grateful for participants in the NHS and HPFS cohorts, US State Cancer Registries (for the full list of states see[1]) and the Odyssey cluster (FAS Division of Science, Research Computing Group at Harvard University.). Funding support by NIH R01 CA49449, P01 CA87969, UM1 CA186107, and UM1 CA167552.

**Leeds**: Funding from Cancer Research UK (project grant C8216/A6129 and programme award C588/A4994) and by the NIH (R01 CA83115). The UK National Cancer Research supported participant recruitment. We specifically acknowledge the support of Patricia Mack, Kate Gamble, Paul King, and Dr Amy Downing.

**QIMR**: We specifically thank Dixie Statham, Ann Eldridge, Marlene Grace, Kerrie McAloney, Natalie Garden, Reshika Chand, Lisa Bowdler, Leanne Wallace, David Smyth, Harry Beeby, and Daniel Park. The Brisbane Twin Nevus Study was supported by Australian National Health and Medical Research Council (NHMRC) grants (241944, 339462, 389927, 389875, 389891, 389892, 389938, 442915, 442981, 496739, 552485, 552498, 1031119), Adult twin data collection by NIH grant (AA011998_5978), over-50 twins by a donation from Mr. George Landers of Chania, Crete. Genotyping was funded by the NHMRC (552498 and 1049894). We also acknowledge the U.S. National Institutes of Health's support through grants AA07535, AA10248, AA13320, AA13321, AA13326, AA14041, and MH66206. This work was also funded by the FP-5 GenomEUtwin Project (QLG2-CT-2002-01254). Genotyping for individuals using the Illumina 370K was funded by an access award to Dr. Richard Todd and performed at the Center for Inherited Disease Research, Baltimore. Nicholas K. Hayward, David C. Whiteman, David L. Duffy, and Grant W. Montgomery were supported by the NHMRC Fellowships scheme during the collection of these samples.

**The Raine Study**: We acknowledge the support for cohort coordination and data collection from the Raine Study and Lions Eye Institute. The Raine Study is supported and funded by The University of Western Australia (UWA), The Telethon Institute for Child Health Research, Raine Medical Research Foundation, UWA Faculty of Medicine, Dentistry and Health Sciences, Women and Infant Research Foundation and Curtin University. Genotyping was funded by the

NHMRC (1021105). Support for the Raine Study examinations and naevi count was provided by the Lions Eye Institute, the Australian Foundation for the Prevention of Blindness, and the Ophthalmic Research Institute of Australia. The Pawsey Supercomputing Centre provided computation resources to carry out genetic analyses with funding from the Australian Government and the Government of Western Australia.

**The Rotterdam Study**: We specifically acknowledge the assistance and support of Ada Hooghart, Corina Brussee, Riet Bernaerts‑Biskop, Patricia van Hilten, Pascal Arp, Jeanette Vergeer, Maarten Kooijman and Lennart Karssen. The Rotterdam study was supported by the Netherlands Organisation of Scientific Research (NWO); Erasmus Medical Center and Erasmus University, Rotterdam, The Netherlands; Netherlands Organization for Health Research and Development (ZonMw); UitZicht; the Research Institute for Diseases in the Elderly; the Ministry of Education, Culture and Science; the Ministry for Health, Welfare and Sports; the European Commission (DG XII); the Municipality of Rotterdam; the Netherlands Genomics Initiative/NWO; Center for Medical Systems Biology of NGI; Stichting Lijf en Leven; Stichting Oogfonds Nederland; Landelijke Stichting voor Blinden en Slechtzienden; Algemene Nederlandse Vereniging ter Voorkoming van Blindheid; Medical Workshop; Heidelberg Engineering; Topcon Europe BV. Research Institute for Diseases in the Elderly (014-93-015; RIDE2), the Netherlands Genomics Initiative (NGI)/Netherlands Organisation for Scientific Research (NWO) project nr. 050-060-810.

F.L. was supported by the Erasmus University Rotterdam (EUR) fellowship and the Chinese recruiting program "The 1000 Talents Plan" for young scholars. GWAS genotyping was funded by the Netherlands Organisation of Scientific Research NWO Investments (nr. 175.010.2005.011, 911-03-012).

**Twins eye study in Tasmania**: This work was supported by an NHMRC Enabling Grant (2004‑2009, 350415, 2005-2007); Clifford Craig Medical Research Trust; Ophthalmic Research Institute of Australia; American Health Assistance Foundation; Peggy and Leslie Cranbourne Foundation; Foundation for Children; Jack Brockhoff Foundation, and the National Eye Institute of the National Institute of Health grants RO1EY01824601 from 2007 to 2010. D. Mackey was supported by a Pfizer Australia Senior Research Fellowship and S. MacGregor was a recipient of an Australian NHMRC Career Development Award. Genotyping was funded by an NHMRC Medical Genomics Grant; US NIH/National Eye Institute (1RO1EY018246), Australian sample imputation analyses were carried out on the Genetic Cluster Computer, which is financially supported by the Netherlands Scientific Organization (NWO48005003).

**TwinsUK**: TwinsUK is funded by the Wellcome Trust, Medical Research Council, Versus Arthritis, European Union Horizon 2020, Chronic Disease Research Foundation (CDRF), Wellcome Leap Dynamic Resilience Programme (co-funded by Temasek Trust), Zoe Ltd, the National Institute for Health and Care Research (NIHR) Clinical Research Network (CRN) and

Biomedical Research Centre based at Guy's and St Thomas' NHS Foundation Trust in partnership with King's College London.

**New nevus samples**:
**QSkin (Phase I and Phase II):** The QSkin study was supported by grants from the National Health and Medical Research Council (NHMRC) of Australia (APP1073898; APP1063061; APP1185416). David Whiteman is supported by an NHMRC Investigator Grant (APP2026567).

**AGDS:** We are forever grateful to the participants for their willingness to participate in our studies. The authors thank our colleagues Richard Parker, Simone Cross, Scott Gordon, and Lenore Sullivan for their valuable work coordinating administrative and operational aspects of the AGDS project. Data collection for AGDS was possible thanks to funding from the Australian National Health & Medical Research Council (NHMRC APP1086683).

**KYAMS:** The Kidskin Young Adult Myopia Study was supported by funding from Perpetual Impact Philanthropy (IPAP2015/0230) and the National Health and Medical Research Council (1121979).

**Melanoma meta-analysis Samples from** [14]

**NCI:** This study was supported by the Intramural Research Program of the Division of Cancer Epidemiology and Genetics, National Cancer Institute (NCI), National Institutes of Health (NIH) and Department of Health and Human Services (DHHS).

**AOCS/OCAC/SEARCH:** AOCS/OCAC/SEARCH is accessible via European Genome–Phenome Archive. We acknowledge their support and data, and the contribution of the study nurses, research assistants and all clinical and scientific collaborators in generation of these data. We also acknowledge their funding sources: OCAC (NIH grant no. U19CA148112), SEARCH team (Cancer Research UK grant no.C490/A16561), AOCS (US Army Medical Research and Material Command under grant no. DAMD17-01-1-0729, The Cancer Council Victoria, Queensland Cancer Fund, The Cancer Council New South Wales, The Cancer Council South Australia, The Cancer Foundation of Western Australia, The Cancer Council Tasmania and the National Health and Medical Research Council of Australia (NHMRC) (grant nos. ID400413 and ID400281, as well as support from S. Boldeman, the Agar family, Ovarian Cancer Action (UK), Ovarian Cancer Australia and the Peter MacCallum Foundation).

**MelaNostrum Consortium:** We thank the participants of the MelaNostrum Consortium from Italy (Genoa, L'Aquila, Rome, Padua, Milan, Florence and Bergamo), Spain (Valencia and Barcelona), Greece (Athens) and Cyprus (Nicosia) who provided data and biospecimens for this study. The Consortium is partially supported by the Intramural Research Program of the Division of Cancer Epidemiology and Genetics, NCI, NIH, DHHS. Funding for the University of Genoa and Genetics of Rare Cancers, Ospedale Policlinico San Martino came from Italian Ministry of Health 5 × 1000 per la Ricerca Corrente to Ospedale Policlinico San Martino and AIRC IG

15460. The research at the Melanoma Unit in Barcelona was supported by the Spanish Fondo de Investigaciones Sanitarias grant nos. PI15/00716 and PI15/00956 cofinanced by FEDER ‘Una manera de hacer Europa’; CIBER de Enfermedades Raras of the Instituto de Salud Carlos III, Spain, cofinanced by European Development Regional Fund ‘A way to achieve Europe’ ERDF; AGAUR 2014_SGR_603 of the Catalan Government, Spain; European Commission, contract no. LSHC-CT-2006-018702 (GenoMEL) and by the European Commission under the 7th Framework Programme, Diagnostics; ‘Fundació La Marató de TV3’ grant no. 201331-30, Catalonia, Spain; ‘Fundación Científica de la Asociación Española Contra el Cáncer’ grant no. GCB15152978SOEN, Spain, and CERCA Programme/Generalitat de Catalunya. Melanoma research at the Department of Dermatology, University of L’Aquila, Italy was supported by the Italian Ministry of the University and Scientific Research (PRIN-2012 grant no. 2012JJX494).

**Q-MEGA/QTWIN:** The Q-MEGA/QTWIN study was supported by the Melanoma Research Alliance, the NIH NCI (grant nos. CA88363, CA83115, CA122838, CA87969, CA055075, CA100264, CA133996 and CA49449), the NHMRC (grant nos. 200071, 241944, 339462, 380385, 389927, 389875, 389891, 389892,389938, 443036, 442915, 442981, 496610, 496675, 496739, 552485, 552498 and APP1049894), the Cancer Councils New South Wales, Victoria and Queensland, the Cancer Institute New South Wales, the Cooperative Research Centre for Discovery of Genes for Common Human Diseases, Cerylid Biosciences (Melbourne), the Australian Cancer Research Foundation, The Wellcome Trust (grant no. WT084766/Z/08/Z) and donations from N. and S. Hawkins. S. MacGregor acknowledges fellowship support from the Australian National Health and Medical Research Council and from the Australian Research Council.

**Endometriosis:** Contributors to the Endometriosis collection: Anjali K. Henders, S.H. Kennedy, S. Macgregor, N.G. Martin, S. Missmer, G.W. Montgomery, D.R. Nyholt, J.N. Painter, S.A. Treloar, L. Wallace, K.T. Zondervan. Acknowledgements: We acknowledge all the participants in the QIMR and endometriosis studies. We thank Anjali Henders, Leanne Wallace, and Lisa Bowdler for project management, sample processing and database development. We thank Endometriosis Associations for supporting study recruitment and S. Nicolaides and the Queensland Medical Laboratory for assistance with blood collection including pro bono collection and delivery of blood samples. Funding: This work was supported by the Cooperative Research Centre (CRC) for Discovery of Genes for Common Human Diseases, Cerylid Biosciences (Melbourne), The Wellcome Trust and donations from Neville and Shirley Hawkins. Endometriosis sample genotyping was funded by a grant from the Wellcome Trust (WT084766/Z/08/Z) and NHMRC (496610, GNT1049472, GNT1050208 ). G.W.M. is supported by the NHMRC Fellowships scheme. D.R.N. was supported by the NHMRC Fellowship (613674) and Australian Research Council (ARC) Future Fellowship (FT0991022) schemes.

**EPIGENE:** We thank melanoma patients, Sullivan and Nicolaides Pathology, Queensland Medical Laboratories, and IQ Pathology for their involvement and support. This cohort was supported by the National Health and Medical Research Council of Australia (APP442960).

**QSkin:** The QSkin Study could not have occurred without the valuable contribution of the Queenslanders who took part, and for that we thank them. The authors gratefully acknowledge the valuable contributions of all staff, students and colleagues who have been associated with the project since its inception. We would like to recognise Australian National Health and Medical Research Council funding (project grant APP106306, programme grant 552429)

**Princess Alexandra Hospital (PAH) samples:** We acknowledge the support and assistance of study participants and Adele Greene (securing funding), Mark Smithers (cohort establishment), as well as Casey Rowe and Maryrose Malt for consenting and collection of patient samples. Funding for the PAH collection was via the University of Queensland Diamantina Institute, the Meehan Foundation, NHMRC CDF (1125290), and Cancer Council Queensland (1125237).

**AMFS:** This work was supported by the National Health and Medical Research Council of Australia (NHMRC) project grants 566946, 107359, 211172, and program grant number 402761 to GJM and RFK. Work was also funded by the Cancer Councils of Victoria, Queensland, and New South Wales (project grants 77/00, 06/10, 371) and by US NIH RO1 grant CA-83115-01A2 and 2R01CA083115-11A1. Anne E. Cust is supported by fellowships from the Cancer Institute NSW and the NHMRC. We gratefully thank the support and involvement of all participants, research coordinators, interviewers, examiners and data management staff.

**Study of Digestive Health:** Controls for use with the Q-MEGA_omni dataset were derived from the Study of Digestive Health group (SDH) which was funded by NCI grant 5 RO1 CA 001833-02. Its contents are solely the responsibility of the authors and do not necessarily represent the official views of the National Cancer Institute. We gratefully acknowledge the cooperation of the following institutions: Sullivan and Nicolaides Pathology (Brisbane); Queensland Medical Laboratory (Brisbane); Queensland Health Pathology Services (Brisbane); Institute of Medical and Veterinary Science (Adelaide); SouthPath (Adelaide). We also acknowledge the contribution of the study nurses and research assistants and would like to thank all of the people who participated in the study. DCW was supported by an NHMRC Research Fellowship (APP1058522).

**Inflammatory Bowel Disease (IBD):** We gratefully acknowledge the aid in identifying study participants by Sullivan and Nicolaides Pathology, Queensland Medical Laboratories and the Queensland Health Pathology Service. The IBD study would like to recognise and thank Peter Schultz, Lauren Aoude, Loralie Parsonson, Stephen Walsh, Mitchell Stark, John Cardinal and Herlina Handoko for technical support.

This work was funded by US NCI grant CA 001833-03. Its contents are solely the responsibility of the authors and do not necessarily represent the official views of the National Cancer Institute. PMW and DCW are Senior Research Fellows of the National Health and Medical Research Council (NHMRC) of Australia. NP was supported by a NHMRC PhD scholarship. The funding bodies played no role in the design or conduct of the study, the collection, management, analysis, or interpretation of the data, or the preparation, review or approval of the manuscript.

**Essen-Heidelberg:** The study was supported by a grant from Deutsche Forshungsgemeinschaft (GZ: SCHA 422/11-1).

**Western Australian Melanoma Health Study (WAMHS):** The WAMHS, and the salaries of its staff and PhD students, was funded by the Scott Kirkbride Melanoma Research Centre. The Cancer Council Western Australia is also acknowledged for current salary support for Sarah Ward (Capacity Building and Collaboration grant). We acknowledge the donation of time and samples from study participants, the WAMHS study team, and the WAMHS Management Committee. This work could not have occurred without the assistance and support of the Western Australian (WA) DNA Bank, and the Ark at The University of WA, and the WA Cancer Registry, for which we are grateful.

**Australian & New Zealand Registry of Advanced Glaucoma (ANZRAG):** Participant collection was funded by the Royal Australian and New Zealand College of Ophthalmology Eye Foundation. Genotyping was supported by NHMRC grants 535074 and 1023911. ANZRAG also acknowledges funding from the BrightFocus Foundation and a Ramaciotti Establishment Grant. The authors acknowledge the support of Ms. Bronwyn Usher‐Ridge in patient recruitment and data collection, and Dr Patrick Danoy and Dr Johanna Hadler for genotyping.

**Generations Scotland:** Generation Scotland received core support from the Chief Scientist Office of the Scottish Government Health Directorates [CZD/16/6] and the Scottish Funding Council [HR03006]. Genotyping of the GS:SFHS samples was carried out by the Genetics Core Laboratory at the Wellcome Trust Clinical Research Facility, Edinburgh, Scotland and was funded by the Medical Research Council UK and the Wellcome Trust (Wellcome Trust Strategic Award "STratifying Resilience and Depression Longitudinally" (STRADL) Reference 104036/Z/14/Z)."

**Michigan:** The authors acknowledge the University of Michigan Precision Health Initiative and Medical School Central Biorepository for providing biospecimen storage, management, processing and distribution services and the Center for Statistical Genetics in the Department of Biostatistics at the School of Public Health for genotype data curation, imputation, and management in support of this research.

**Brisbane Nevus Morphology Study:** This work was funded by project grant numbers NHMRC 1004999, 1062935 and the Centre of Research Excellence for the Study of Naevi 1099021. This research was carried out at the Translational Research Institute, Woolloongabba, QLD 4102, Australia.

The Translational Research Institute is supported by a grant from the Australian Government

**MIA:** This work was supported by research funding from Melanoma Institute Australia, the National Health and Medical Research Council of Australia (NHMRC) through program grants to GJM, RAS & GVL and from Cancer Institute New South Wales and infrastructure grants from Macquarie University and the Australian Cancer Research Foundation. R.A.S. and G.V.L. are supported by NHMRC Fellowships, and G.V.L. is supported by the University of Sydney Medical Foundation. RS is supported by the Melanoma Institute Australia (MIA), the New South Wales Department of Health, NSW Health Pathology, the National Health and Medical Research Council of Australia (NHMRC) and Cancer Institute NSW and reports receiving fees for professional services from Merck Sharp & Dohme, GlaxoSmithKline Australia, Bristol-Myers Squibb, Dermpedia, Novartis Pharmaceuticals Australia Pty Ltd, Myriad, NeraCare GmbH and Amgen.

**MDACC:** We thank the individuals who volunteered to participate in this project. This work was supported by the National Cancer Institute of the National Institutes of Health through SPORE grant P50 CA093459 and Cancer Center Support Grant P30 CA016672 (Clinical Trials Support Resource), as well as by philanthropic contributions to The University of Texas MD Anderson Cancer Center Moon Shots Program, The University of Texas MD Anderson Cancer Center Various Donors Melanoma and Skin Cancers Priority Program Fund, the Miriam and Jim Mulva Research Fund, the McCarthy Skin Cancer Research Fund, and the Marit Peterson Fund for Melanoma Research.

**Genoa:** The Genoa study was supported by The Italian Ministry of Health Grant RF-2016-02362288 and 5x1000 per la ricerca corrente. The authors acknowledge Dr. William Bruno for his continuous work on melanoma families.

**Barcelona:** The research at the Melanoma Unit in Barcelona is partially funded by Spanish Fondo de Investigaciones Sanitarias grants PI15/00716, PI15/00956, PI18/00419 and PI18/01077; CIBER de Enfermedades Raras of the Instituto de Salud Carlos III, Spain, co-financed by European Development Regional Fund "A way to achieve Europe" ERDF; AGAUR 2017_SGR_1134 of the Catalan Government, Spain; European Commission under the 6th Framework Programme, Contract No. LSHC-CT-2006-018702 (GenoMEL) and by the European Commission under the 7th Framework Programme, Diagnoptics; The National Cancer Institute (NCI) of the US National Institute of Health (NIH) (CA83115); a grant from "Fundació La Marató de TV3" 201331-30, Catalonia, Spain; a grant from "Fundación Científica de la Asociación Española Contra el Cáncer" GCB15152978SOEN, Spain, and CERCA Programme / Generalitat de Catalunya. Part of the work was carried out at the Esther Koplowitz Center, Barcelona.

**ICR:** We thank Breast Cancer Now and the Institute of Cancer Research for funding and acknowledge National Health Service funding to the Royal Marsden NHS Foundation Trust and Institute of Cancer Research NIHR Biomedical Research Centre.

**MELARISK, France:** This work was supported by grants from Institut National du Cancer (INCa-PL016 and INCa_5982) to FD, Ligue Nationale Contre Le Cancer (PRE 09/FD) to FD, Programme Hospitalier de Recherche Clinique (AOM-07-195) to MFA and FD, Ministère de l'Enseignement Supérieur et de la Recherche and Institut National du Cancer (INCa) to GML. MB was supported by fellowships from Ligue Nationale Contre Le Cancer and Fondation pour la Recherche Médicale (FDT20130928343). The authors thank the French Family study group for contributing data to the MELARISK study3,5,64, the Supplementation in Vitamins and Mineral Antioxidants (SU.VI.MAX) study group for giving access to data of the Su.VI.MAX study65 and the Epidemiological Study on the Genetics and Environment of Asthma (EGEA) cooperative group for giving access to data of the EGEA study (https://egeanet.vjf.inserm.fr). We acknowledge that the biological specimens of the French MELARISK study were obtained from the Institut Gustave Roussy and Fondation Jean Dausset–CEPH Biobanks.
